# Why Americans Choose Raw Milk: Evidence from a National Quantitative Survey

**DOI:** 10.64898/2026.10.26.26366487

**Authors:** Sophia Ruser, Cristina Rojo, Katherine Koebel, Calvin Slaughter, Ece Bulut, Renata Ivanek

**Affiliations:** College of Veterinary Medicine, Cornell University, S1-026 Schurman Hall, Ithaca, NY 14853; Yale School of Medicine, 333 Cedar Street, New Haven, CT 06510; College of Agriculture and Life Sciences, Cornell University, Stocking Hall, Ithaca, NY, 14853

**Keywords:** Unpasteurized milk, consumer behavior, risk perception, food safety, heating

## Abstract

Raw milk consumption is a controllable source of foodborne pathogen exposure in the United States (US), yet existing national estimates of raw milk consumption behaviors are outdated and do not capture emerging factors such as consumers’ trusted information sources, sociopolitical influences, and at-home risk mitigation behaviors. We conducted a nationally representative cross-sectional survey of n=1078 US adults to estimate the prevalence of raw milk consumption and at-home thermal treatment practices and identify predictors of these behaviors. Multivariable logistic regression models were used to evaluate predictors of raw milk consumption and thermal treatment behavior using adjusted odds ratios (aORs). Among 1,075 eligible respondents, 15% consumed raw milk in the last six months, including 5% exclusively and 10% in addition to pasteurized milk. In multivariable analysis, raw milk consumption was independently associated with younger age, cohabitation with high-risk individuals for foodborne infections (including children ≤5 years of age, and elderly, pregnant or immunocompromised individuals) who had consumed raw milk, and perceptions of raw milk taste and safety. Adults aged ≥55 years had lower odds of consumption than those aged 18–24 years (aOR, 0.05; 95% CI, 0.02–0.15), while those living with a high-risk household member who consumed raw milk had higher odds of consumption (aOR, 4.39; 95% CI, 2.69–7.22). Perceiving raw milk to have higher risk of causing illness was associated with lower odds of raw milk consumption (aOR: 0.32, 95% CI: 0.17-0.62) compared to believing the risk to be equal across milk types, and thermal treatment was reported by 38% of exclusive raw milk consumers (n=58) and 47% of consumers of both milk types (n=106). Raw milk consumption in the US appears to be substantially more common than previously reported and shaped by factors such as taste, safety, and household raw milk consumption among high-risk individuals for foodborne infections. The proportion of raw milk consumers already practicing at-home thermal treatment suggests a potential opportunity for risk mitigation.

## Introduction

Raw milk, or milk that has not undergone pasteurization, is a known vehicle of foodborne exposure with pathogens such as *Salmonella* spp., pathogenic *Escherichia coli*, *Campylobacter* spp., *Staphylococcus aureus*, *Brucella* spp., *Coxiella burnetii*, and *Listeria monocytogenes* ^1^. In the United States (US), human consumption of raw dairy products is associated with 840 times more illnesses and 45 times more hospitalizations compared to consumption of pasteurized dairy products when adjusted for servings consumed ^2^. Multiple outbreak investigations have linked raw milk consumption to hospitalization and death, including 75 outbreaks with 675 illnesses during 2013–2018 alone ^1, 3, 4^. In the US, raw milk access varies by state, with regulations ranging from prohibitions on sale to allowances for retail or on-farm sales, cow– or herd-share programs, and sales labeled for animal consumption ^5^.

Despite these risks, a 2016 nationally representative survey estimated that ∼4–5% of US adults consume raw milk annually ^6^. Consumers are primarily younger adults, rural residents, and individuals in states with legal sale access ^6, 7^. Concerningly, a substantial proportion of consumers report living with individuals at high risk for foodborne infections, including children and older adults ^6, 7^. Beyond demographic patterns, raw milk consumption is shaped by beliefs about perceived health and safety benefits, many of which are not supported by scientific evidence. Proponents of raw milk consumption claim that it is more nutritious than pasteurized milk and may reduce lactose intolerance, allergies, and asthma ^8^. Prior studies demonstrate that perceived benefits often undermine established danger in decision-making, with many consumers denying illness risk and expressing low trust in public health authorities ^9^. Other motivating factors for raw milk consumption include taste, a desire to support local agriculture, and strong social influences such as family traditions and peer networks ^9, 10^. Online platforms have facilitated the dissemination of claims promoting the purported health benefits of raw milk ^10^. More broadly, social media has contributed to the rapid spread of nutritional misinformation, potentially influencing consumer food choices and perceptions of risk ^11^. Consumer preferences for natural and minimally processed foods may also contribute to the appeal of raw milk ^12^.

In addition to individual motivations, raw milk consumption is shaped by its legal availability, which varies substantially across the US. The transport of raw milk for human consumption between states is prohibited under federal regulation, as outlined in the Pasteurized Milk Ordinance ^13^. However, state-level regulations range from complete prohibition of sales to more permissive guidelines, including the allowance of sales labeled for animal consumption, cow– or herd-share programs, and on-farm or retail sales ^5^. These regulations govern commercial access but do not necessarily prohibit consumption; for example, some states permit dairy producers to consume raw milk from their own animals ^14^.

Regardless of how raw milk is obtained, thermal treatment before consumption represents a potential means of reducing pathogen risk. The most common industrial pasteurization method for fluid milk in the US is high temperature short time (HTST) pasteurization, which involves heating milk to 72°C and holding it for 15 seconds, yielding >5-log reductions of pathogens of concern while preserving its nutritional value ^15, 16^. Other common methods include low temperature long time (LTLT), ultrahigh temperature (UHT), and vat pasteurization (Vat-PT) ^13^. Historically, when commercially pasteurized milk was less widely available, home boiling was promoted as a practical public health intervention to reduce milk-borne disease. Although guidance for home boiling remains available today, thermal treatment prevalence among contemporary US raw milk consumers remains unknown, despite its investigation as a risk-reduction strategy in populations such as Italy and Ethiopia ^17–21^. Understanding this behavior may be particularly important; qualitative research suggests many US raw milk consumers place greater trust in their own food safety practices and familiar producers than in industrial processing methods or government guidance ^22^.

Despite these insights, critical gaps remain in understanding contemporary raw milk consumption patterns in the US. Previous estimates are now dated and provide limited insight into emerging influences, impacting public health messaging. We sought to address these knowledge gaps through a quantitative, nationally representative survey with the following objectives: to determine the prevalence of raw milk consumption in the US, estimate the frequency of at-home raw milk heating among raw milk consumers, and identify predictors of these behaviors.

## Methods

### Study Population and Questionnaire Survey Administration

The questionnaire survey was developed to evaluate consumer perceptions and behaviors regarding raw and pasteurized milk. The target population consisted of adults (i.e., aged ≥18 years) residing in the US. The survey was administered by Qualtrics Research Services (QRS) ^23^ from February 14–17, 2025, with a target sample size of 1,000 participants. QRS used demographic quotas for sex, age, geographic region, and education level to approximate the US adult population. The full questionnaire is provided in Supplementary Table S1, and additional details regarding QRS recruitment and sampling procedures are provided in Supplementary Text S1. Cornell Institutional Review Board (IRB) approved the study protocol as an exemption on January 23, 2025 (protocol number IRB0149147).

### Variable Definition and Outcome Modeling

Table 1 defines the variables used in the statistical analyses and their corresponding survey questions. Whether a respondent had consumed raw milk in the past six months (Q1) and at-home thermal treatment behavior among raw milk consumers (Q2; presented only to respondents who in Q1 responded “Raw dairy milk” or “Both raw and pasteurized dairy milk”) were defined as the primary and secondary outcomes of interest, respectively. Perceptions of taste (Q5), nutrition beliefs (Q6), illness risk (Q7), cohabitation with high-risk individuals for foodborne illness who have consumed raw milk in the past six months (Q4), and H5N1-related milk safety concern (Q8), as well as trusted information source (Q3), were defined as predictors of interest. The H5N1 milk safety question was included to capture how the newly emerged influenza A(H5N1) clade 2.3.4.4b avian influenza (bird flu) outbreak in dairy cattle, ongoing during survey administration, may have affected respondents’ raw milk perceptions and behaviors. Age (Q9), sex at birth (Q10), geographic region (Q11), education level (Q12), and political orientation (Q13) were assessed as potential confounders (Table 1).

**Table 1.** Variable Definitions, Roles in Statistical Analysis, and Corresponding Survey Questions.

|  |  |  |  |
| --- | --- | --- | --- |
| Role | Variable | Definition | Survey Question<sup>1</sup> |
| Primary Outcome | Any Raw Milk Consumption (Model R<sub>Any</sub>) | Binary: Individuals who reported consuming raw milk in any capacity in the last 6 months (including those who consumed raw milk exclusively or in addition to pasteurized milk) compared to those who consumed pasteurized milk only (reference category) | Q1 |
| Primary Outcome | Exclusive Raw Milk Consumption (Model R<sub>Exclusive</sub>) | Binary: Individuals who exclusively consumed raw milk in the last 6 months compared to those who consumed both raw and pasteurized milk or only pasteurized milk (reference category) | Q1 |
| Secondary Outcome | At-Home Thermal Treatment before Any Raw Milk Consumption (Model T<sub>Any</sub>) | Binary: Among any raw milk consumers, individuals who reported heating raw milk more than half the time or always (consistent thermal treatment) compared to those who reported heating half the time or less, i.e., never, less than half the time, or approximately half the time (reference category). "Prefer not to answer" was coded as missing and excluded from analysis. | Q2<sup>2</sup> |
| Secondary Outcome | At-Home Thermal Treatment before Exclusive Raw Milk Consumption (Model T<sub>Exclusive</sub>) | Binary: Among exclusive raw milk consumers, individuals who reported heating raw milk more than half the time or always (consistent thermal treatment) compared to those who reported heating half the time or less, i.e., never, less than half the time, or approximately half the time (reference category). "Prefer not to answer" was coded as missing and excluded from analysis. | Q2<sup>2</sup> |
| Predictor | Trusted Information Source | Categorical: Most trusted source of information regarding milk safety. Response options: "Doctors or nurses" (reference category), "Friends or family," "Farmers," "Government officials," "Social media influencers," "Professors or researchers," "Healer, chiropractor, naturopath," "Other [specify]." Respondents who selected "Prefer not to answer" were categorized as "Uncertain/No response". | Q3 |
| Predictor | Cohabitation with high-risk individuals who have consumed raw milk | Binary: Presence of a high-risk household member (children aged ≤5, adults aged ≥65, individuals with limited immunity, or pregnant individuals) who consumed raw milk in the last 6 months, compared to no at-risk household member (reference category). "Prefer not to answer" responses were grouped with the reference category. | Q4<sup>3</sup> |
| Predictor | Perceived Taste | Categorical: Belief about how the taste of raw milk compares to pasteurized milk. Response options: "Raw is better," "Pasteurized is better," "They are equal" (reference category), with "I don't know" and "Prefer not to answer" grouped as "Uncertain/No response". | Q5 |
| Predictor | Nutrition Beliefs | Categorical: Belief about the nutritional value of raw vs. pasteurized milk. Response options: "Raw is better," "Pasteurized is better," "They are equal" (reference category), with "I don't know" and "Prefer not to answer" grouped as "Uncertain/No response". | Q6 |
| Predictor | Perceived Illness Risk | Categorical: Belief about which type of milk is more likely to cause illness. Response options: "Raw milk," "Pasteurized milk," "They are equal" (reference category), with "I don't know" and "Prefer not to answer" grouped as "Uncertain/No response". | Q7 |
| Predictor | H5N1 Milk Safety Concern | Categorical: Belief about whether consumers should worry about milk safety during a bird flu (H5N1) outbreak in dairy cattle. Response options: "Yes, if consuming raw milk," "Yes, if consuming pasteurized milk," "Yes, if consuming either raw or pasteurized milk", and "No" (reference category), with "I don't know", and "Prefer not to answer" grouped as "Uncertain/No response". | Q8 |
| Potential Confounder | Age Group | Categorical: Self-reported age: 18–24 (reference category), 25–34, 35–44, 45–54, and 55+ (combining 55–64 and 65 or over). | Q9 |
| Potential Confounder | Sex at Birth | Categorical: Sex assigned at birth, Male compared to Female (reference category). Respondents who selected another option or declined to answer were coded as missing and excluded from models including this variable. | Q10 |
| Potential Confounder | Geographic Region | Categorical: US Census region of current residence, derived from state of residence, categorized as Midwest (reference category), Northeast, South, and West. | Q11 |
| Potential Confounder | Education Level | Categorical: Highest level of educational attainment, collapsed into four categories: "High school, equivalent, or less" (reference category), "Some college or technical," "College graduate," and "Graduate or professional." Respondents who declined to answer were coded as missing and excluded from models including this variable. | Q12 |
| Potential Confounder | Political Orientation | Categorical: Self-described political party orientation, collapsed into three categories: "Democrat" (including Democrat orientation qualified as strong, not very strong, or independent-leaning; reference category), "Republican" (including Republican orientations qualified as strong, not very strong, or independent-leaning), and "Independent/Other" (identifying as independent leaning toward neither party or reporting another party affiliation). Respondents who declined to answer were coded as missing and excluded from models including this variable. | Q13 |
| Note: The full survey instrument is available in the Supplemental Materials. (Supplementary Table S1)<br><sup>1</sup>Q = Survey Question number.<br><sup>2</sup>Presented only to respondents who reported consuming raw milk in Q1 (either "Raw dairy milk" or "Both raw and pasteurized dairy milk").<br><sup>3</sup>High-risk household members in Q4, defined as children aged ≤5 years, adults aged ≥65 years, immunocompromised individuals, or pregnant individuals. |  |  |  |

The primary outcome (raw milk consumption; Q1) was dichotomized into two versions to differentiate between distinct consumption patterns: Model R_Any_ for any raw milk consumption (including those who consumed raw milk exclusively or in addition to pasteurized milk) and Model R_Exclusive_ for exclusive raw milk consumption in the last 6 months (Table 1) and identify their predictors.

The secondary outcome, at-home thermal treatment of raw milk, was assessed by asking how frequently respondents heated raw milk (e.g., by boiling or steaming) prior to consumption (Q2). Responses were dichotomized based on consistency of heating: respondents reporting heating more than half the time or always were categorized as consistent heaters, and those reporting heating half the time or less (never, less than half the time, or approximately half the time) served as the reference group. This threshold was selected because infrequent heating still results in regular consumption of unheated raw milk. Respondents who selected “Prefer not to answer” were coded as missing. Because this question applied only to raw milk consumers, the analysis was restricted to respondents reporting raw milk consumption in the past six months and was conducted separately among all raw milk consumers (Model T_Any_; including those who consumed raw milk exclusively or in addition to pasteurized milk) and among exclusive raw milk consumers (Model T_Exclusive_).

### Predictor Variables Preparation and Recoding

For perceived taste (Q5), nutrition beliefs (Q6), perceived illness risk (Q7), and H5N1 safety concern (Q8), responses of “I don’t know,” and “Prefer not to answer” were combined into a single “Uncertain/No response” category, as these responses accounted for a large proportion of the sample for these questions, particularly Q5 and Q6. For Q5, Q6, and Q7, “They are equal” was retained as a distinct response category, as it represented a substantive belief about each question rather than uncertainty, while “I don’t know” and “Prefer not to answer” were grouped together, representing a lack of any strong opinion. For trusted information source (Q3), respondents who selected “prefer not to answer” or were also categorized as “Uncertain/No response” category as well.

Cohabitation with pre-defined high-risk individuals for foodborne illness within the household who consumed raw milk in the last 6 months (Q4) was assessed with a multiple-select question. For analysis, responses were used to ascertain counts of each high-risk subgroup for descriptive purposes. Additionally, a binary indicator for cohabitation with a high-risk individual who consumed raw milk was created to identify respondents whose household included at least one high-risk individual (i.e., children aged 5 or younger, adults aged 65 or older, immunocompromised individuals, or pregnant individuals) who had consumed raw milk in the past six months, compared to those who indicated that this did not apply to any member of their household (Table 1). Respondents who declined to answer this question (n=24) were grouped with those reporting no cohabitation with a high-risk individual who consumed raw milk for multivariable analysis.

Age (Q9) was collapsed from six categories into five, with respondents aged 55–64 and 65 or over combined into a single “55 or older” group. Sex at birth (Q10) was analyzed as reported; respondents who selected any option aside from male or female or declined to answer (n = 3) were coded as missing and excluded from models including this variable. Geographic region (Q11) was derived from self-reported state of residence and categorized according to U.S. Census Bureau regions ^24^ (Table 1). Education level (Q12) was collapsed into four categories: “high school or less” (below high school, some high school, high school graduate or GED; reference category); “some college or technical” (technical/trade/vocational school, some college or two-year associate degree); “college graduate” (four-year degree); and “graduate or professional” (post-graduate or professional training). Political orientation (Q13) was collapsed into three categories: respondents identifying as strong Democrat, not very strong Democrat, or independent leaning Democrat were classified as “Democrat”; those identifying as strong Republican, not very strong Republican, or independent leaning Republican were classified as “Republican”; and those identifying as independent leaning toward neither party or reporting another party affiliation were classified as “Independent/Other.” Respondents who declined to answer the education or political orientation questions were coded as missing and excluded from models including this variable.

### Statistical Analysis

All statistical analyses were performed using R. To verify the national representativeness of the sample, we compared the demographic characteristics of the survey participants with the 2020 US Census data ^25^ using a one-sample z test. Variables included in these comparisons were geographic region, age, sex, and education level.

In statistical analyses of associations for two versions of the primary outcome (Models R_Any_ and R_Exclusive_), we first performed univariable logistic regression between each predictor and each of the outcomes of interest. Odds ratios (ORs) and the associated Wald 95% confidence intervals (CIs) were calculated as measures of association. To ensure potential confounders were not excluded prematurely, all variables with a p-value < 0.10 were carried over into multivariable analysis.

For the two versions of the secondary outcome (Models T_Any_ and T_Exclusive_), associations between candidate predictors and thermal treatment were first assessed using the original five-level heating frequency response, prior to dichotomization. Fisher’s exact tests were used in place of chi-square tests due to small expected cell counts. Predictors were then examined using the dichotomized outcome following the same screening and model selection procedure described above (i.e., heating always or more than half the time vs approximately half the time or less frequently/never), and analyses were repeated under two alternative dichotomization thresholds (heating at least half the time vs less than half the time or never; any frequency of heating versus never) to assess whether findings depended on a particular categorization. Similarly to the univariable analysis of the primary outcome, all variables with a p-value < 0.10 were carried over into multivariable analysis, and corresponding multivariable models were constructed.

### Multivariable Model Selection and Validation

Final model selection for an outcome of interest was performed using forward stepwise selection based on the Akaike Information Criterion (AIC). For each final model, two-way interactions between significant predictors were evaluated after variable selection, followed by the Benjamini-Hochberg corrections for multiple comparisons of the interaction terms. An adjusted Odds Ratio (aOR) was used as a measure of the association between a specific predictor and an outcome, while controlling for other variables in the final model. Model performance for the primary outcomes was assessed using the Hosmer-Lemeshow goodness-of-fit test and the Area Under the Receiver Operating Characteristic Curve (AUC-ROC). Statistical significance was set at p < 0.05.

## Results

### Study Population

The final number of responses was 1,078, as QRS received more responses than anticipated before closing the survey. The responses were determined to be nationally representative relative to the 2020 US Census with respect to geographic region (p=0.91), sex (p=0.99), age (p=0.23), and level of education (p=0.43) using one-sample z tests. After excluding respondents who did not meet eligibility criteria (i.e., under 18 (n = 3) or not US residents (n = 0)), there were 1,075 remaining eligible responses (Table 2). Those who either declined to answer the question regarding milk consumption or indicated that they did not consume any dairy milk (n = 177) were removed. This yielded a final analytical sample of 898 respondents evaluated in the univariable and multivariable statistical analysis.

**Table 2.**
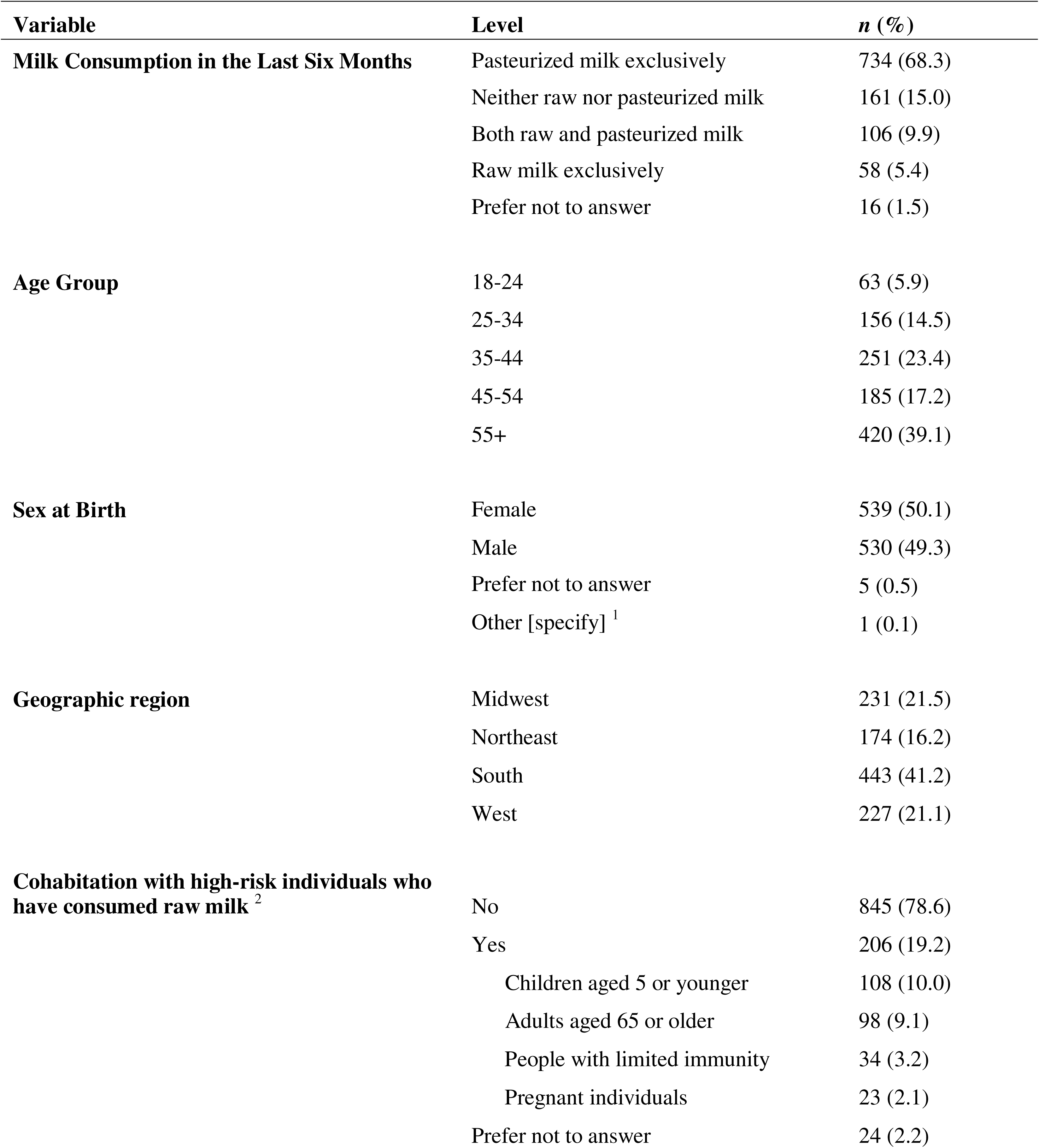
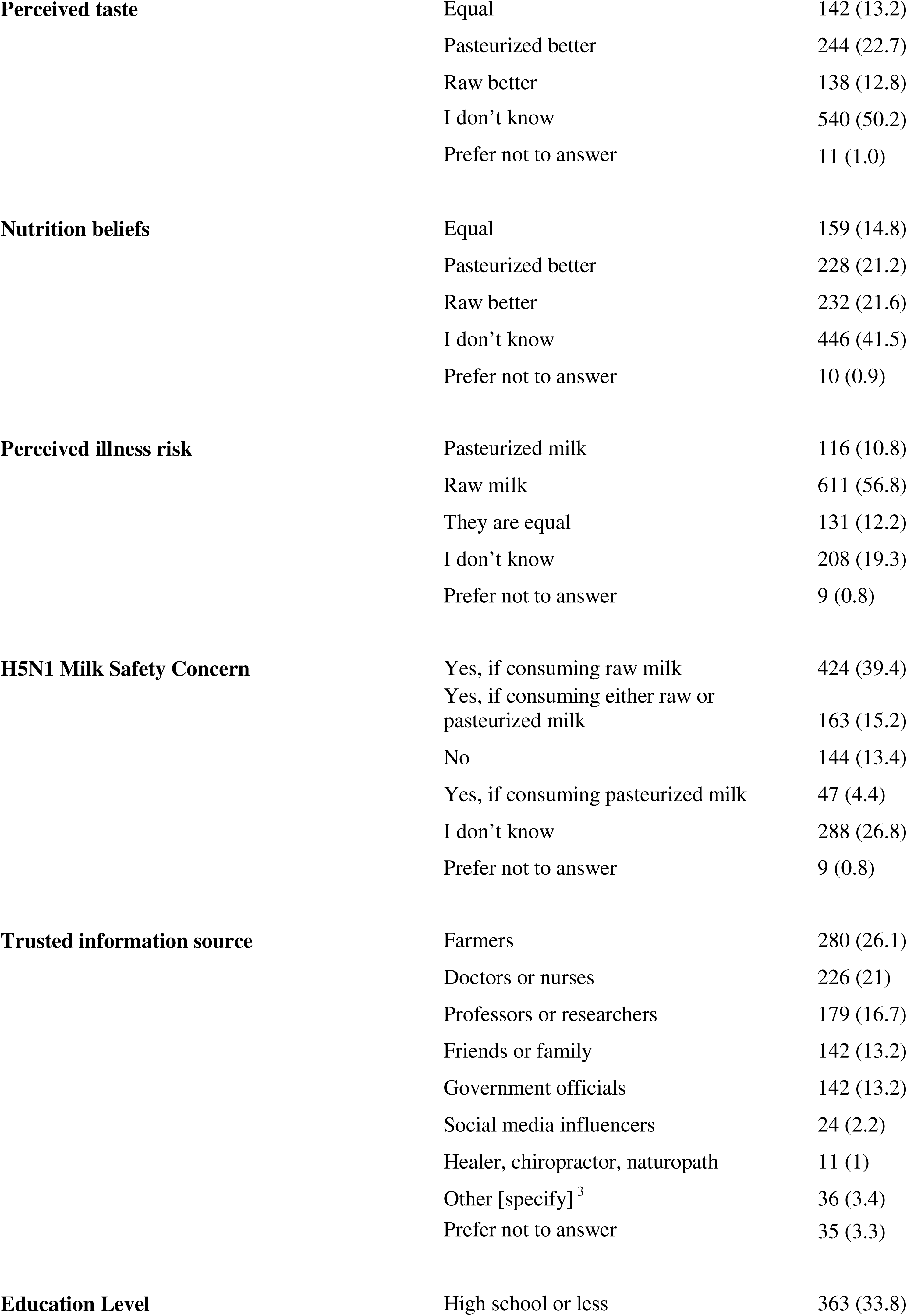
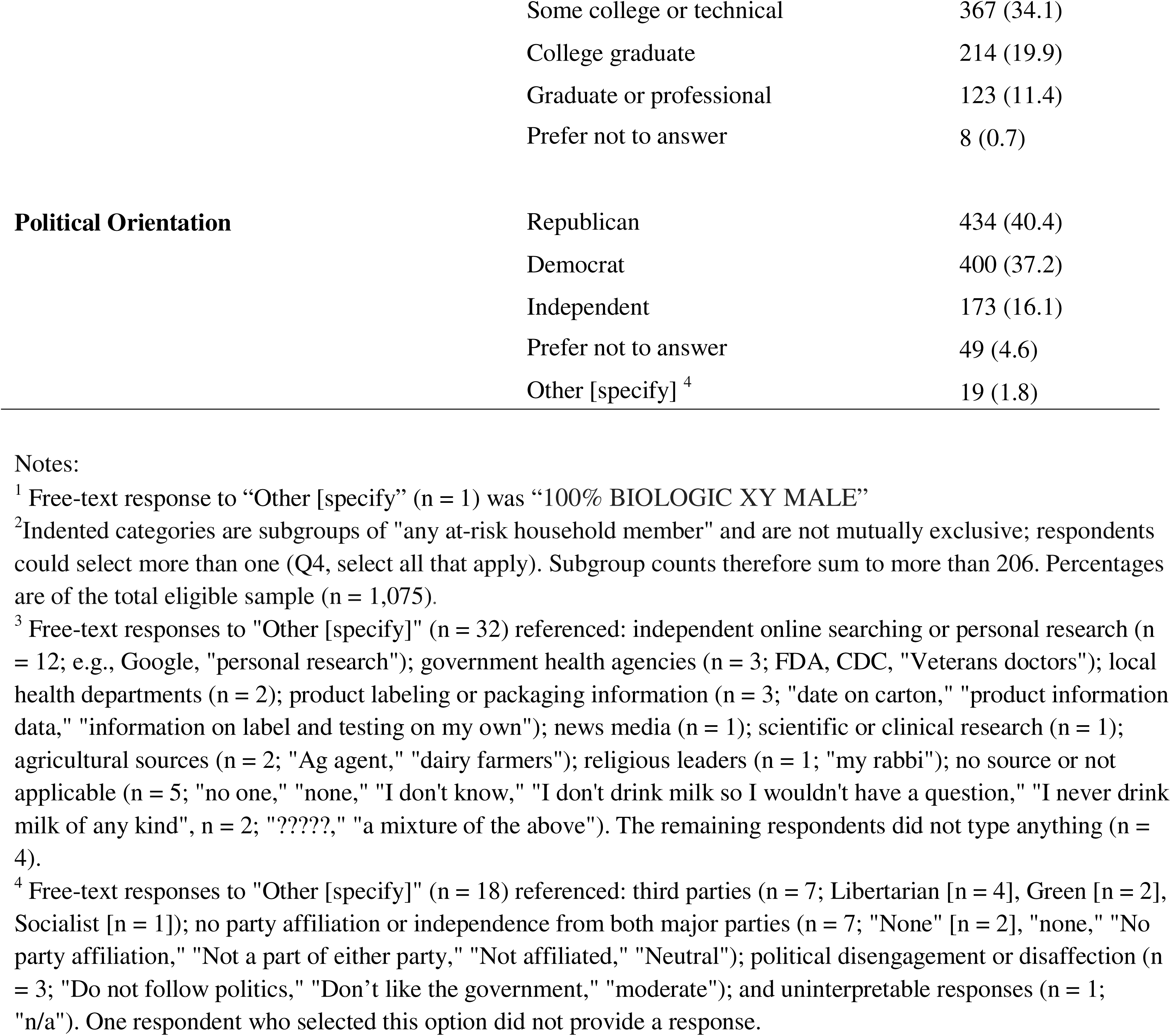
Demographic characteristics, consumption behaviors and perceptions of eligible survey participants (n = 1,075)

| Variable | Level | n (%) |
| --- | --- | --- |
| **Milk Consumption in the Last Six Months** | Pasteurized milk exclusively | 734 (68.3) |
|  | Neither raw nor pasteurized milk | 161 (15.0) |
|  | Both raw and pasteurized milk | 106 (9.9) |
|  | Raw milk exclusively | 58 (5.4) |
|  | Prefer not to answer | 16 (1.5) |
| **Age Group** | 18-24 | 63 (5.9) |
|  | 25-34 | 156 (14.5) |
|  | 35-44 | 251 (23.4) |
|  | 45-54 | 185 (17.2) |
|  | 55+ | 420 (39.1) |
| **Sex at Birth** | Female | 539 (50.1) |
|  | Male | 530 (49.3) |
|  | Prefer not to answer | 5 (0.5) |
|  | Other [specify] <sup>1</sup> | 1 (0.1) |
| **Geographic region** | Midwest | 231 (21.5) |
|  | Northeast | 174 (16.2) |
|  | South | 443 (41.2) |
|  | West | 227 (21.1) |
| **Cohabitation with high-risk individuals who have consumed raw milk <sup>2</sup>** | No | 845 (78.6) |
|  | Yes | 206 (19.2) |
|  | Children aged 5 or younger | 108 (10.0) |
|  | Adults aged 65 or older | 98 (9.1) |
|  | People with limited immunity | 34 (3.2) |
|  | Pregnant individuals | 23 (2.1) |
|  | Prefer not to answer | 24 (2.2) |
| **Perceived taste** | Equal | 142 (13.2) |
|  | Pasteurized better | 244 (22.7) |
|  | Raw better | 138 (12.8) |
|  | I don't know | 540 (50.2) |
|  | Prefer not to answer | 11 (1.0) |
| **Nutrition beliefs** | Equal | 159 (14.8) |
|  | Pasteurized better | 228 (21.2) |
|  | Raw better | 232 (21.6) |
|  | I don't know | 446 (41.5) |
|  | Prefer not to answer | 10 (0.9) |
| **Perceived illness risk** | Pasteurized milk | 116 (10.8) |
|  | Raw milk | 611 (56.8) |
|  | They are equal | 131 (12.2) |
|  | I don't know | 208 (19.3) |
|  | Prefer not to answer | 9 (0.8) |
| **H5N1 Milk Safety Concern** | Yes, if consuming raw milk | 424 (39.4) |
|  | Yes, if consuming either raw or pasteurized milk | 163 (15.2) |
|  | No | 144 (13.4) |
|  | Yes, if consuming pasteurized milk | 47 (4.4) |
|  | I don't know | 288 (26.8) |
|  | Prefer not to answer | 9 (0.8) |
| **Trusted information source** | Farmers | 280 (26.1) |
|  | Doctors or nurses | 226 (21) |
|  | Professors or researchers | 179 (16.7) |
|  | Friends or family | 142 (13.2) |
|  | Government officials | 142 (13.2) |
|  | Social media influencers | 24 (2.2) |
|  | Healer, chiropractor, naturopath | 11 (1) |
|  | Other [specify] <sup>3</sup> | 36 (3.4) |
|  | Prefer not to answer | 35 (3.3) |
| **Education Level** | High school or less | 363 (33.8) |
|  | Some college or technical | 367 (34.1) |
|  | College graduate | 214 (19.9) |
|  | Graduate or professional | 123 (11.4) |
|  | Prefer not to answer | 8 (0.7) |
| **Political Orientation** |  |  |
|  | Republican | 434 (40.4) |
|  | Democrat | 400 (37.2) |
|  | Independent | 173 (16.1) |
|  | Prefer not to answer | 49 (4.6) |
|  | Other [specify] <sup>4</sup> | 19 (1.8) |

Notes:
<sup>1</sup> Free-text response to "Other [specify]" (n = 1) was "100% BIOLOGIC XY MALE"
<sup>2</sup> Indented categories are subgroups of "any at-risk household member" and are not mutually exclusive; respondents could select more than one (Q4, select all that apply). Subgroup counts therefore sum to more than 206. Percentages are of the total eligible sample (n = 1,075).
<sup>3</sup> Free-text responses to "Other [specify]" (n = 32) referenced: independent online searching or personal research (n = 12; e.g., Google, "personal research"); government health agencies (n = 3; FDA, CDC, "Veterans doctors"); local health departments (n = 2); product labeling or packaging information (n = 3; "date on carton," "product information data," "information on label and testing on my own"); news media (n = 1); scientific or clinical research (n = 1); agricultural sources (n = 2; "Ag agent," "dairy farmers"); religious leaders (n = 1; "my rabbi"); no source or not applicable (n = 5; "no one," "none," "I don't know," "I don't drink milk so I wouldn't have a question," "I never drink milk of any kind", n = 2; "?????", "a mixture of the above"). The remaining respondents did not type anything (n = 4).
<sup>4</sup> Free-text responses to "Other [specify]" (n = 18) referenced: third parties (n = 7; Libertarian [n = 4], Green [n = 2], Socialist [n = 1]); no party affiliation or independence from both major parties (n = 7; "None" [n = 2], "none," "No party affiliation," "Not a part of either party," "Not affiliated," "Neutral"); political disengagement or disaffection (n = 3; "Do not follow politics," "Don't like the government," "moderate"); and uninterpretable responses (n = 1; "n/a"). One respondent who selected this option did not provide a response.

### Descriptive Statistics

Of the eligible respondents (n=1,075), most reported consuming only pasteurized milk (68%) in the past six months, while 15% reported not consuming dairy milk at all and were therefore excluded from further analysis. In this sample, 10% of respondents consumed raw milk alongside pasteurized milk, while 5% consumed only raw milk (Table 2). Thermal treatment of raw milk (e.g., by microwave, boiling, or steaming) was practiced to some extent by a large portion of raw milk consumers, with 87% of raw and pasteurized milk drinkers reporting heating “Less than half the time” or more, and 71% of exclusive raw milk consumers doing so (Table 3). Those who consumed a combination of raw and pasteurized milk were more likely to practice at-home heating of raw milk overall, with responses concentrated in the intermediate frequency categories (47% reported heating “More than half the time” or “Always”). Exclusive consumers were more likely to report never heating their raw milk (29% vs. 13% of non-exclusive consumers), while both exclusive and non-exclusive consumers selected “Always” at a very similar frequency of about one-quarter.

**Table 3.**
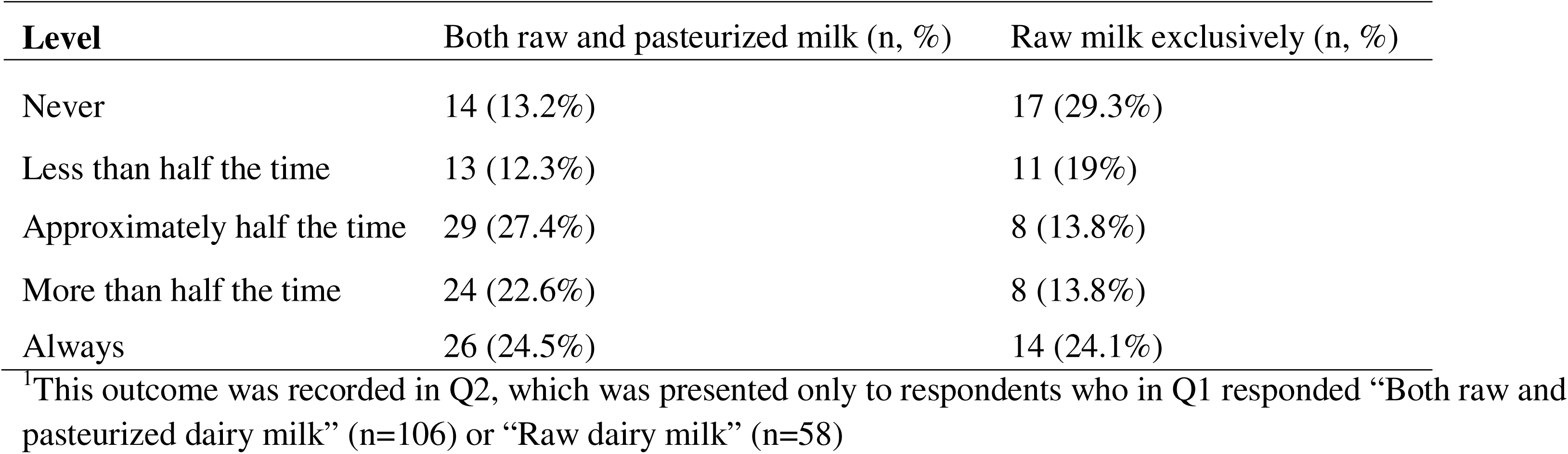
At-Home Thermal Treatment Frequency by Consumption (outcome in Models T_Any_ and T_Exclusive_)^1^.

| Level | Both raw and pasteurized milk (n, %) | Raw milk exclusively (n, %) |
| --- | --- | --- |
| Never | 14 (13.2%) | 17 (29.3%) |
| Less than half the time | 13 (12.3%) | 11 (19%) |
| Approximately half the time | 29 (27.4%) | 8 (13.8%) |
| More than half the time | 24 (22.6%) | 8 (13.8%) |
| Always | 26 (24.5%) | 14 (24.1%) |

<sup>1</sup> This outcome was recorded in Q2, which was presented only to respondents who in Q1 responded "Both raw and pasteurized dairy milk" (n=106) or "Raw dairy milk" (n=58)

## Raw Milk (Models R)

### Univariable analysis

#### Predictors of Any Raw Milk Consumption (Model R_Any_)

Univariable logistic regression on the analytical sample of 898 participants revealed several statistically significant demographic and perception-related associations with any raw milk consumption (either raw only or both pasteurized and raw) (Table 4). Males showed slightly higher odds of consumption than females (OR: 1.58, 95% CI: 1.12-2.24). Geographically, residents in the Northeast and South were each almost twice as likely to have consumed both raw and pasteurized milk compared to those in the Midwest (p < 0.05 for both).

**Table 4.**
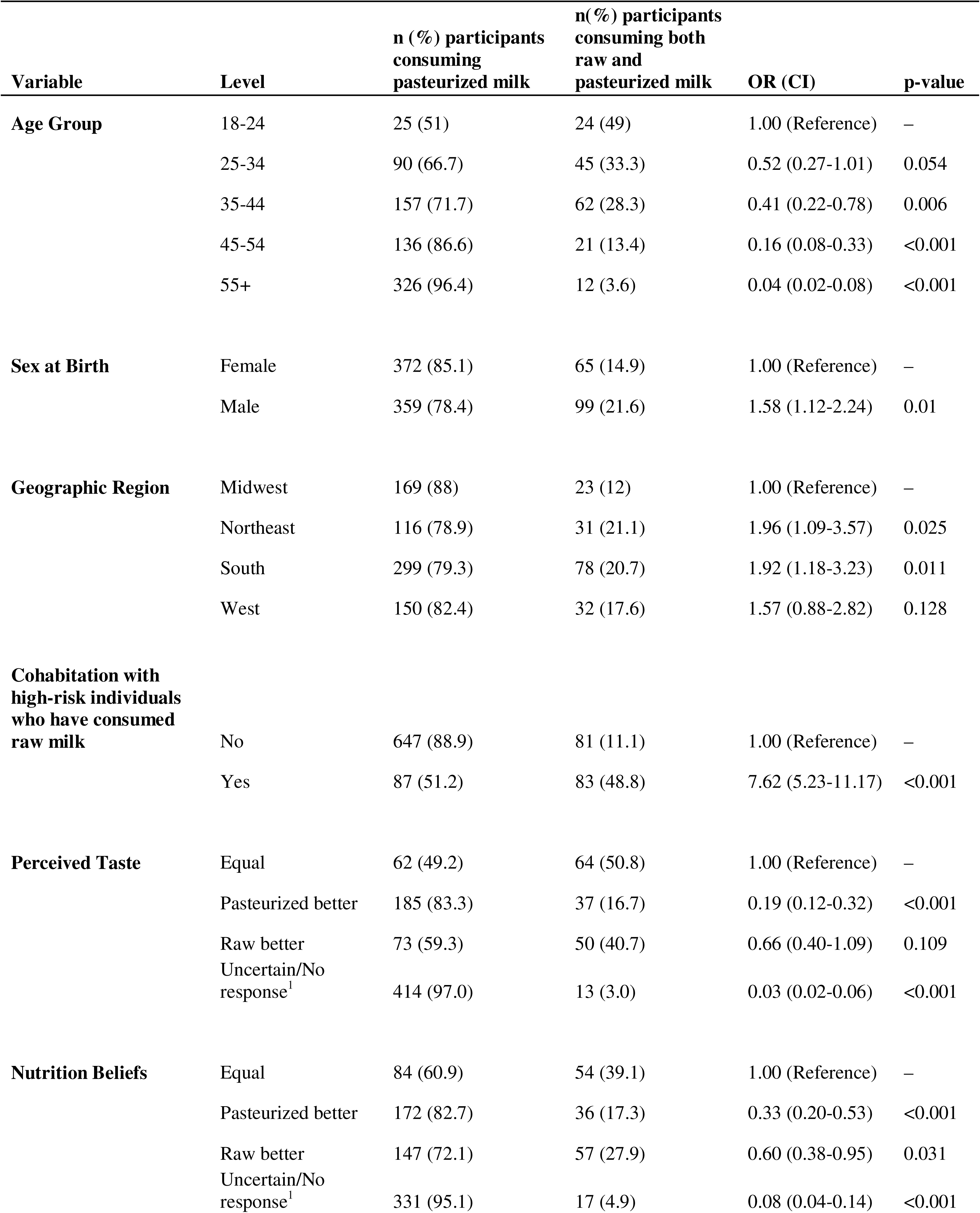
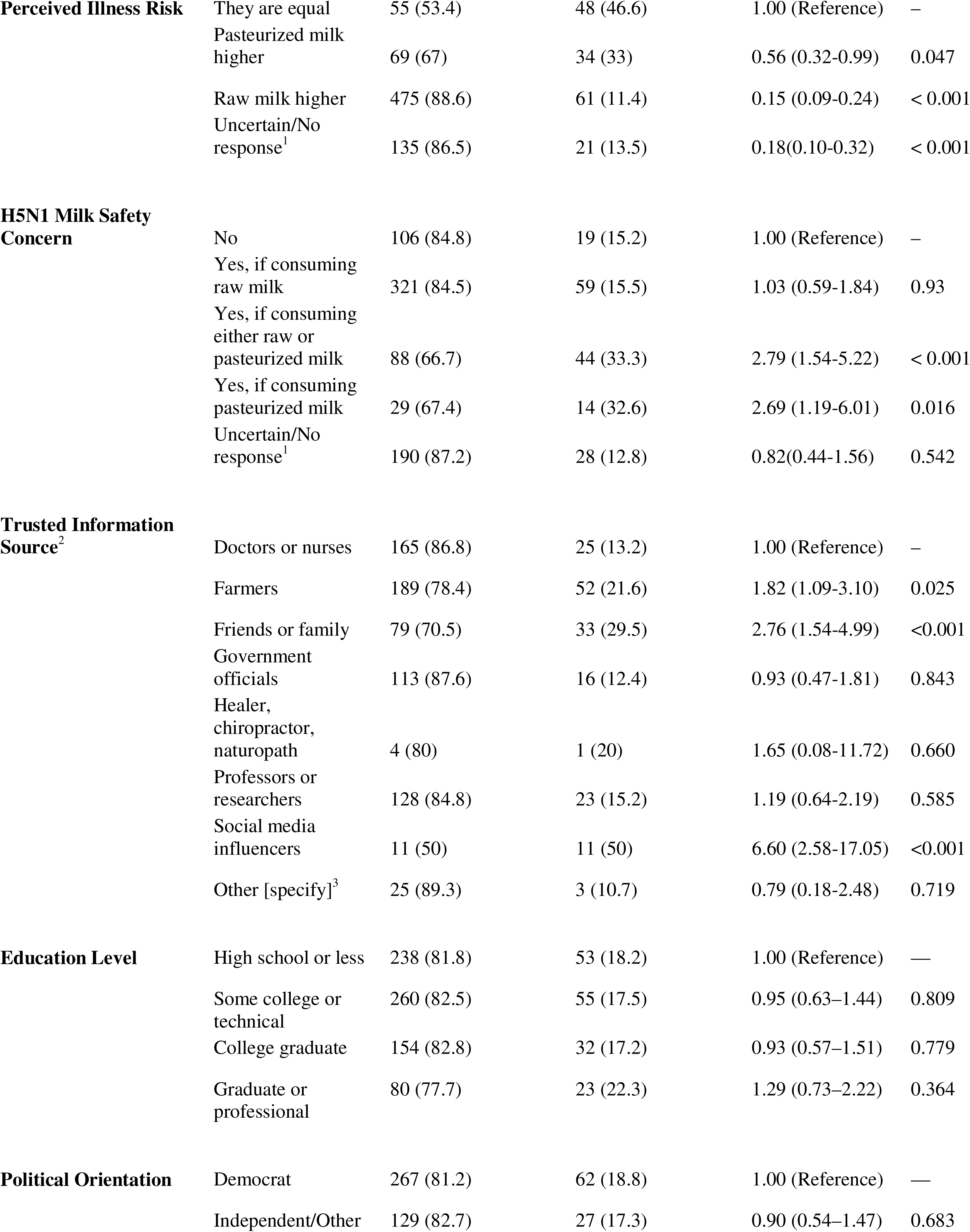
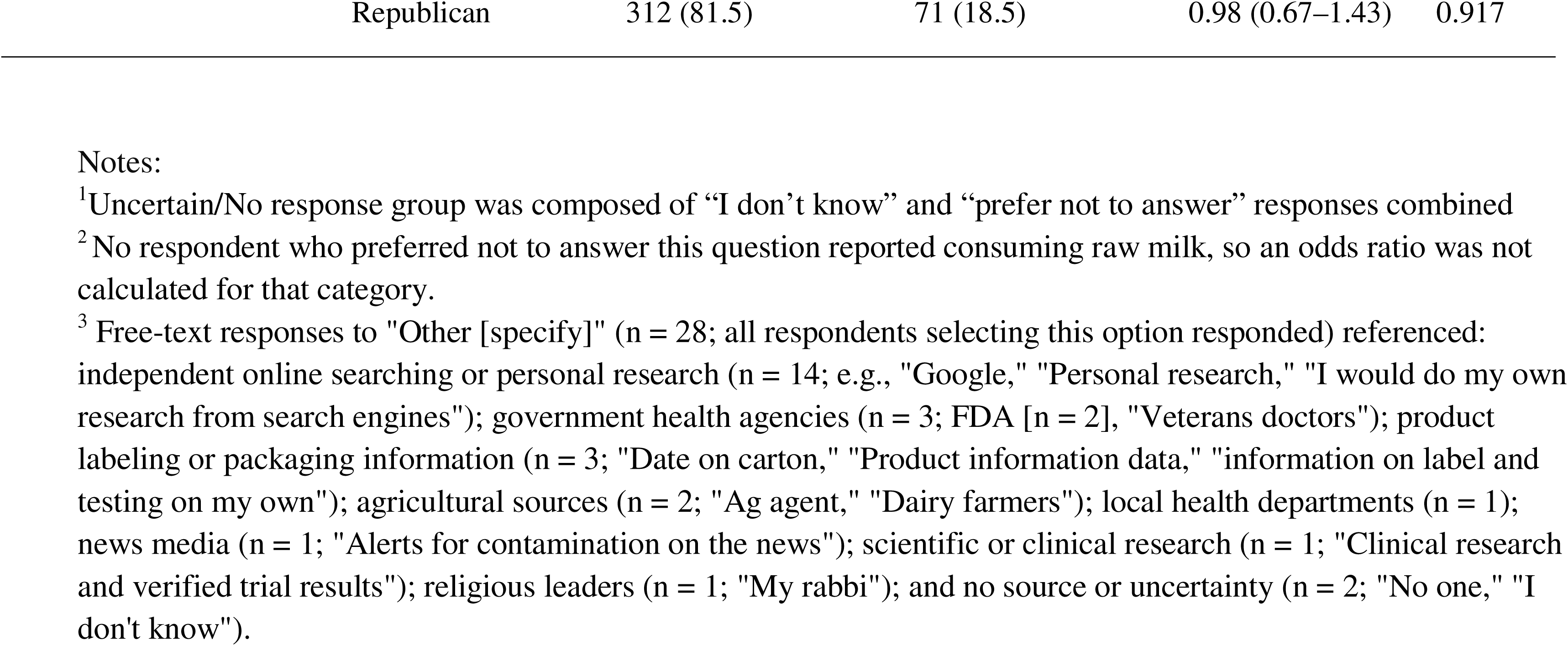
Univariable Association of Demographic and Perceptual Factors with Any Raw Milk Consumption (Model R_Any_, baseline: consuming pasteurized milk only; n=898)

Compared with respondents who viewed raw and pasteurized milk as nutritionally “Equal”, those who selected “Pasteurized better” (OR 0.33, 95% CI 0.20–0.53) or “Raw better” (OR 0.60, 95% CI 0.38–0.95) had lower odds of consuming both raw and pasteurized milk. For taste, respondents who indicated “Pasteurized better” had significantly lower odds of consuming both raw and pasteurized milk (OR 0.19, 95% CI 0.12–0.32), as did those who were grouped as “Uncertain/No response” (OR: 0.03, CI: 0.02-0.06) (Table 4). Regarding perception of H5N1 risk, respondents concerned about both, selecting “Yes, if consuming either raw or pasteurized milk” (OR: 2.79, 95% CI: 1.54–5.22) or “Yes, if consuming pasteurized milk” (OR: 2.69, 95% CI: 1.19–6.01) had significantly higher odds of consuming both raw and pasteurized milk compared to those indicating “No” concern, while concern specific to raw milk (“Yes, if consuming raw milk”) was not significantly associated with consumption (p = 0.930). Perceived illness risk was also influential; compared to believing “They are equal”, all other selections (“Pasteurized higher [risk]”, “Raw milk higher [risk]”, or “Uncertain/no response”) showed significantly lower odds of consumption. Trusted information source was a significant predictor of consumption as well. Compared to those indicating the most trust in “Doctors or nurses”, those listening to “Social media influencers”, “Friends or family”, and “Farmers” all showed significantly higher odds of consumption, while “Government officials” and “Professors or researchers” were not significantly associated with consumption (Table 4). Cohabitation with high-risk individuals who had consumed raw milk in the past six months was one of the strongest predictors in the univariable analysis; participants answering “Yes” to living with at-risk members who consumed raw milk had significantly higher odds of consumption themselves (OR: 7.62, 95% CI: 5.23–11.17, p < 0.001).

#### Predictors of Exclusive Raw Milk Consumption (Model R_Exclusive_)

In univariable logistic regression to identify predictors associated with exclusive raw milk consumption (Table 5), many predictors aligned with the results for Model R_Any_, with a few important differences. Age remained a significant predictor for exclusive consumption (p < 0.001), but the protective effect of the older age group was slightly less pronounced than in Model R_Any_. Respondents over the age of 55 were still the least likely group to consume raw milk exclusively (OR: 0.05, 95% CI: 0.01–0.13) compared to the 18–24 reference group. While sex and geographic region were significant predictors of raw milk consumption in Model R_Any_, they did not show a statistically significant association with exclusive raw milk consumption.

**Table 5.** Univariable Association of Demographic and Perceptual Factors with Exclusive Raw Milk Consumption (Model R_Exclusive_, baseline: consuming pasteurized milk only or both raw and pasteurized milk; n=898)

| Variable | Level | n(%) for participants consuming raw and pasteurized milk | n(%) for participants exclusively consuming raw milk | OR (CI) | p-value |
| --- | --- | --- | --- | --- | --- |
| Age Group | 18-24 | 37 (75.5) | 12 (24.5) | 1.00 (Reference) | – |
|  | 25-34 | 121 (89.6) | 14 (10.4) | 0.36 (0.15-0.85) | 0.018 |
|  | 35-44 | 201 (91.8) | 18 (8.2) | 0.28 (0.12-0.63) | 0.002 |
|  | 45-54 | 148 (94.3) | 9 (5.7) | 0.19 (0.07-0.48) | <0.001 |
|  | 55+ | 333 (98.5) | 5 (1.5) | 0.05 (0.01-0.13) | <0.001 |
| Sex at Birth | Female | 409 (93.6) | 28 (6.4) | 1.00 (Reference) | – |
|  | Male | 428 (93.4) | 30 (6.6) | 1.02 (0.60-1.75) | 0.931 |
| Geographic Region | Midwest | 184 (95.8) | 8 (4.2) | 1.00 (Reference) | – |
|  | Northeast | 140 (95.2) | 7 (4.8) | 1.15 (0.39-3.28) | 0.792 |
|  | South | 349 (92.6) | 28 (7.4) | 1.85 (0.86-4.41) | 0.136 |
|  | West | 167 (91.8) | 15 (8.2) | 2.07 (0.87-5.25) | 0.107 |
| **Cohabitation with a high-risk individual who has consumed raw milk** |  |  |  |  |  |
|  | No | 706 (97.0) | 22 (3.0) | 1.00 (Reference) | – |
|  | Yes | 134(78.8) | 36(21.2) | 8.62 (4.96-15.32) | <0.001 |
| **Perceived Taste** |  |  |  |  |  |
|  | Equal | 114 (90.5) | 12 (9.5) | 1.00 (Reference) | – |
|  | Pasteurized better | 211 (95) | 11 (5) | 0.50 (0.21-1.16) | 0.105 |
|  | Raw better | 94 (76.4) | 29 (23.6) | 2.93 (1.45-6.26) | 0.004 |
|  | Uncertain/No response<sup>1</sup> | 421 (98.6) | 6 (1.4) | 0.14 (0.05-0.36) | < 0.001 |
| **Nutrition Beliefs** |  |  |  |  |  |
|  | Equal | 125 (90.6) | 13 (9.4) | 1.00 (Reference) | – |
|  | Pasteurized better | 196 (94.2) | 12 (5.8) | 0.59 (0.26-1.34) | 0.203 |
|  | Raw better | 179 (87.7) | 25 (12.3) | 1.34 (0.67-2.80) | 0.414 |
|  | Uncertain/No response<sup>1</sup> | 340 (97.7) | 8 (2.3) | 0.23 (0.09-0.55) | 0.001 |
| **Perceived Illness Risk** |  |  |  |  |  |
|  | They are equal | 92 (89.3) | 11 (10.7) | 1.00 (reference) | – |
|  | Pasteurized milk | 90 (87.4) | 13 (12.6) | 1.21 (0.51-2.89) | 0.664 |
|  | Raw milk | 512 (95.5) | 24 (4.5) | 0.39 (0.19-0.86) | 0.014 |
|  | Uncertain/No response<sup>1</sup> | 146 (93.6) | 10 (6.4) | 0.57(0.23-1.41) | 0.223 |
| **H5N1 Milk Safety Concern** |  |  |  |  |  |
|  | No | 119 (95.2) | 6 (4.8) | 1.00 (reference) | – |
|  | Yes, if consuming either raw or pasteurized milk | 121 (91.7) | 11 (8.3) | 1.80 (0.66-5.38) | 0.260 |
|  | Yes, if consuming pasteurized milk | 38 (88.4) | 5 (11.6) | 2.61 (0.72–9.14) | 0.130 |
|  | Yes, if consuming raw milk | 354(93.2) | 26 (6.8) | 1.46 (0.62-3.99) | 0.419 |
|  | Uncertain/No response<sup>1</sup> | 208 (95.4) | 10 (4.6) | 0.95 (0.35–2.86) | 0.928 |
| **Trusted Information Source<sup>2</sup>** | Doctors or nurses | 181 (95.3) | 9 (4.7) | 1.00 (Reference) | — |
|  | Farmers | 223 (92.5) | 18 (7.5) | 1.62 (0.73-3.87) | 0.249 |
|  | Friends or family | 96 (85.7) | 16 (14.3) | 3.35 (1.46-8.19) | 0.005 |
|  | Government officials | 122 (94.6) | 7 (5.4) | 1.15 (0.40-3.18) | 0.782 |
|  | Professors or researchers | 149 (98.7) | 2 (1.3) | 0.27 (0.04-1.07) | 0.097 |
|  | Social media influencers | 18 (81.8) | 4 (18.2) | 4.47 (1.12-15.29) | 0.021 |
|  | Healer, chiropractor, naturopath | 4 (80.0) | 1 (20.0) | 5.03 (0.24-38.71) | 0.167 |
|  | Other [specify]<sup>3</sup> | 27 (96.4) | 1 (3.6) | 0.74 (0.04-4.20) | 0.784 |
| **Education Level** | High school or less | 266 (91.4) | 25 (8.6) | 1.00 (Reference) | — |
|  | Some college or technical | 297 (94.3) | 18 (5.7) | 0.64 (0.34–1.20) | 0.171 |
|  | College graduate | 176 (94.6) | 10 (5.4) | 0.60 (0.27–1.25) | 0.193 |
|  | Graduate or professional | 99 (96.1) | 4 (3.9) | 0.43 (0.12–1.14) | 0.126 |
| **Political Orientation** | Democrat | 305 (92.7) | 24 (7.3) | 1.00 (Reference) | — |
|  | Independent/Other | 147 (94.2) | 9 (5.8) | 0.78 (0.33–1.66) | 0.534 |
|  | Republican | 361 (94.3) | 22 (5.7) | 0.77 (0.42–1.41) | 0.402 |

Notes:
<sup>1</sup>Uncertain/No response groups “I don’t know” and “prefer not to answer” together
<sup>2</sup>No respondent who preferred not to answer this question reported consuming raw milk, so an odds ratio was not calculated for that category.
<sup>3</sup>Free-text responses to “Other [specify]” (n = 28; all respondents selecting this option provided a response) referenced: independent online searching or personal research (n = 14; e.g., “Google,” “Personal research,” “I would do my own research from search engines”); government health agencies (n = 3; FDA [n = 2], “Veterans doctors”); product labeling or packaging information (n = 3; “Date on carton,” “Product information data,” “information on label and testing on my own”); agricultural sources (n = 2; “Ag agent,” “Dairy farmers”); local health departments (n = 1); news media (n = 1; “Alerts for contamination on the news”); scientific or clinical research (n = 1; “Clinical research and verified trial results”); religious leaders (n = 1; “My rabbi”); and no source or uncertainty (n = 2; “No one,” “I don’t know”).

Cohabitation with a high-risk individual who had consumed raw milk remained a strong predictor of exclusive raw milk consumption; those cohabiting with such an individual had significantly higher odds of exclusive consumption compared to those who did not (OR: 8.62, 95% CI: 4.96–15.32). Taste perception remained a significant motivating factor, with those believing raw milk tasted better having significantly higher odds of exclusive consumption compared to those perceiving the taste as equal (OR: 2.93, 95% CI: 1.45–6.26), while those in the “Uncertain/no response” taste preference category had significantly lower odds (OR: 0.14, 95% CI: 0.05–0.36). For nutrition beliefs, only the “Uncertain/No response” category differed significantly from the reference (OR: 0.23, 95% CI: 0.09–0.55). Perceived illness risk was significant only for those believing raw milk posed greater risk, with that group having lower odds of exclusive consumption (OR: 0.39, 95% CI: 0.19–0.86), and unlike in Model R_Any_, H5N1 safety concern was not a significant predictor of exclusive consumption at any level. Trusted information source continued to be an influential predictor in this univariable analysis. Friends and family and social media influencers remained significant predictors compared to medical professionals, with higher odds of consumption, while farmers, which were significant in Model R_Any_, were not significant predictors of exclusive consumption.

## Multivariable Analysis

Multivariable logistic regression models were constructed for both Model R_Any_ and Model R_Exclusive_ of the main outcome (raw milk consumption) to identify independent predictors of behavior.

### Model R_Any_: Any Raw Milk Consumption

The final multivariable model for Model R_Any_ demonstrated adequate calibration and fit, as validated by the Hosmer-Lemeshow goodness-of-fit test ( ^2^ = 3.137 df = 8, p = 0.926), and excellent predictive discrimination, AUC-ROC of 0.912. In this multivariable Model R_Any_ (Figure 1), several factors remained significant predictors after controlling for other variables. Cohabitation with high-risk individuals who have consumed raw milk was a strong independent predictor; respondents who reported cohabitation with a high-risk individual who consumed raw milk had 4.39 times higher odds of consuming raw milk themselves compared to those who did not (95% CI: 2.69–7.22). Perceptions of taste were also strong predictors of any raw milk consumption. With taste being equal as the reference category, respondents who believed pasteurized milk tasted better had significantly lower odds of consumption (aOR: 0.22, 95% CI: 0.11–0.40), as did those with Uncertain/No response taste preferences (aOR: 0.04, 95% CI: 0.02–0.10). Believing that raw milk tasted better was not significantly associated with consumption.

**Figure 1.**
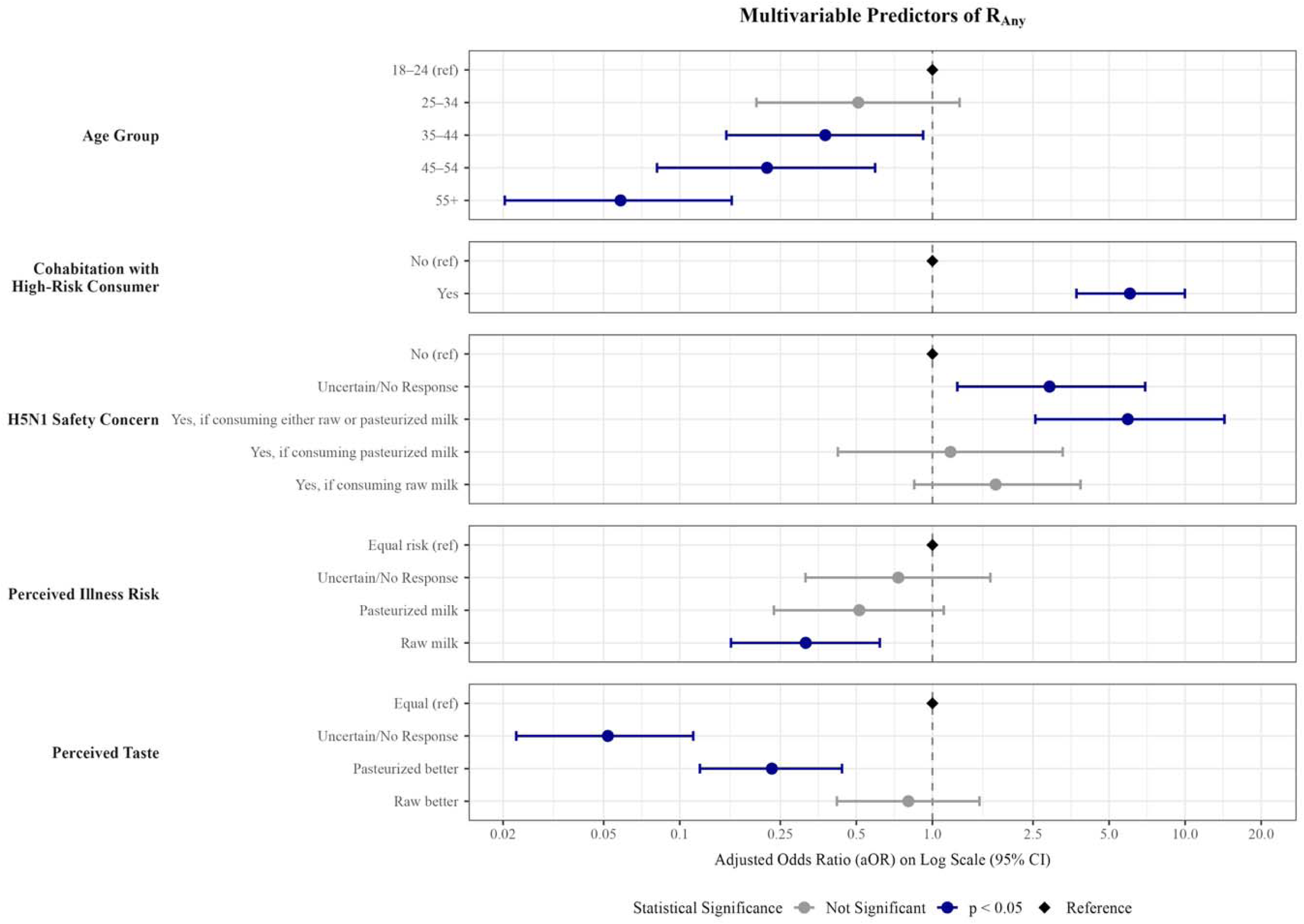
Multivariable predictors of any raw milk consumption (model R_Any_) in the past six months (n = 898)

**Figure 2.**
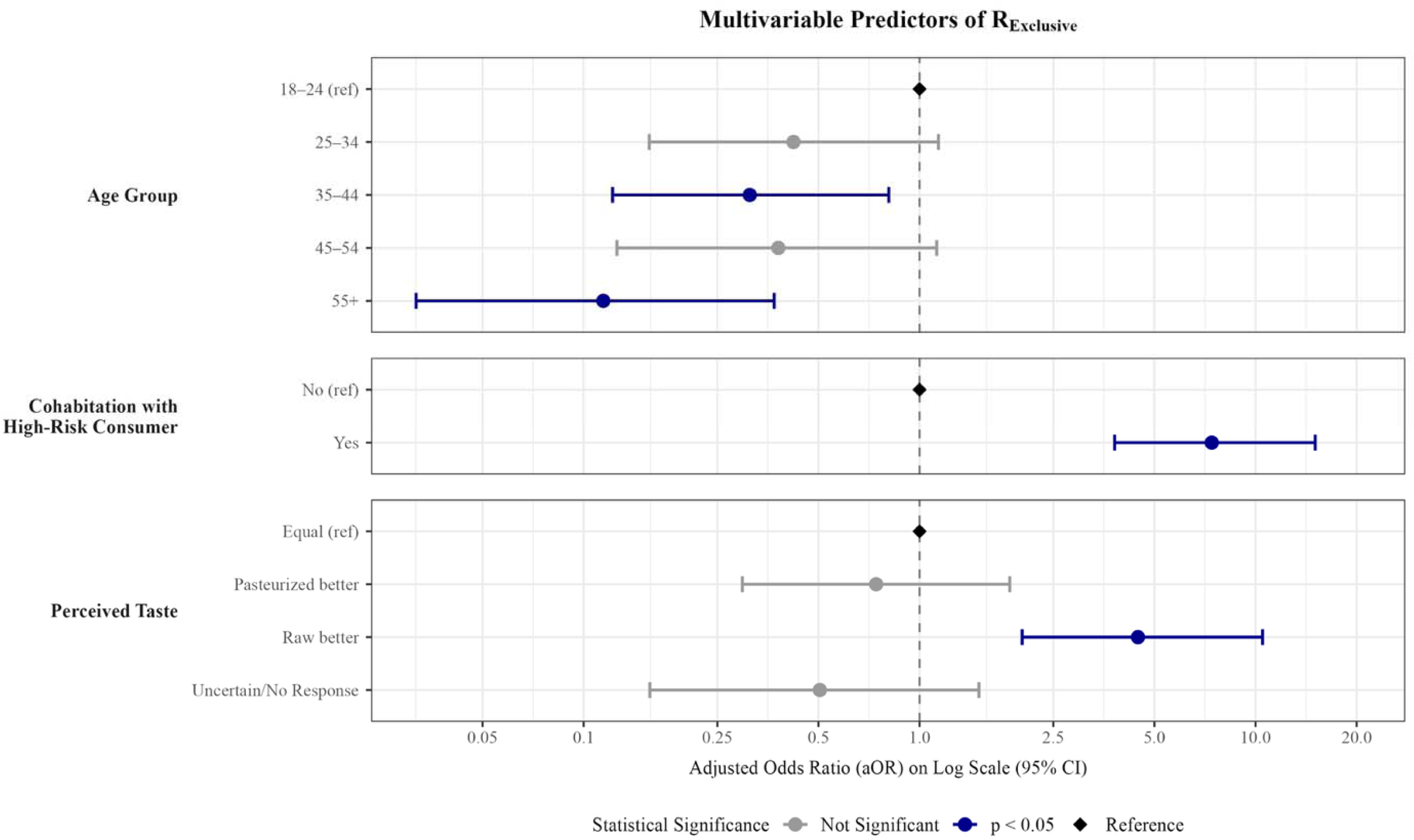
Multivariable predictors of exclusive raw milk consumption (model R_Exclusive_) in the past six months (n = 898)

Safety concerns regarding H5N1 were also influential. Using “No” as the reference, respondents who answered “Yes, if consuming either raw or pasteurized milk” had the highest odds of any raw milk consumption (aOR: 6.60, 95% CI: 2.90–15.62), followed by those in the “Uncertain/No response” group (aOR: 2.67, 95% CI: 1.17–6.27). Perceived illness risk showed significant prediction influence as well; respondents who believed raw milk posed a greater illness risk had lower odds of any raw milk consumption compared to those who believed the risk was equal between milk types (aOR: 0.32, 95% CI: 0.17–0.62). Age remained inversely associated with consumption, with the 55 or older age group showing the lowest odds compared to the 18–24 reference group (aOR: 0.05, 95% CI: 0.02–0.15).

The tests of two-way interactions in this final model identified a significant interaction between taste and cohabitation with high-risk individuals who consumed raw milk in likelihood ratio testing (χ² = 9.38, df = 3, p = 0.026). However, it was one of ten interactions assessed and did not remain significant under the Benjamini-Hochberg correction for multiple testing (both adjusted p = 0.256). While the AIC marginally favored the model including the interaction, given the added complexity of the model, the limited size of the compared groups, difficulties in the interpretation of the “Uncertain/No response” category in this model and the fact that this interaction was not confirmed in Model R_Exclusive_, the interaction was not retained in the final model R_Any_ and is reported as an exploratory finding in supplementary materials (Supplementary Text S2 and Figure S1).

### Model R_Exclusive_: Exclusive Raw Milk Consumption

The final multivariable model for exclusive raw milk showed a similar pattern of predictors. Cohabitation with a high-risk individual who consumed raw milk remained the dominant predictor, with those respondents having 7.41 times higher odds of being exclusive raw milk consumers compared to those not cohabitating with a high-risk individual who consumed raw milk (95% CI: 3.81–15.06). Taste perception was also strongly associated. Using “Equal” as the reference, respondents who believed raw milk tasted better had significantly higher odds of exclusive consumption (aOR: 4.47, 95% CI: 2.02-10.49). Age remained inversely associated with exclusive consumption, with the 55+ group showing the lowest odds compared to the 18–24 reference group (aOR: 0.11, 95% CI: 0.03–0.37). H5N1 safety concern and perceived illness risk were evaluated as candidate predictors but were not retained in the final model, as their inclusion did not improve model fit. We evaluated the model for 2-way interactions but identified none. This model displayed good discrimination, with an AUC of 0.879, and the Hosmer-Lemeshow test supported that it was an adequate fit (p = 0.723).

### Thermal Treatment (Models T)

#### Univariable and Multivariable Analysis

We examined the secondary outcome, at-home thermal treatment, among participants who reported consuming raw milk in the past six months. Thermal treatment was assessed by asking how frequently respondents heated raw milk (e.g., by **boiling** or steaming) prior to consumption (Q2). Heating frequency differed significantly between those who consume both raw and pasteurized milk and exclusive raw milk consumers (Fisher’s exact p = 0.03), driven primarily by the proportion reporting no thermal treatment: 29% (n = 17) of exclusive raw milk consumers reported never heating their milk, compared to 13% (n = 14) of those consuming both raw and pasteurized milk. Additionally, among participants who consumed both raw and pasteurized milk (n = 106), 47% (n = 50) reported heating raw milk more than half the time or always, compared to 38% (n = 22) of those consuming exclusively raw milk (n = 58).

Associations between candidate predictors and thermal treatment were first assessed using the original five-level heating frequency response, before any dichotomization. Among all raw milk consumers (Model T_Any;_ (n=164), comprised of those who consume both raw and pasteurized milk (n=106) and those who drink raw milk exclusively (n=58), with the description of each model being more comprehensively described in supplementary table S2), education level showed the strongest association (p = 0.008) followed by taste (p = 0.013); nutritional beliefs did not reach significance at the 5% threshold. Among exclusive raw milk consumers (Model T_Exclusive_; n=58), perceived illness risk was the only predictor associated with heating frequency (p = 0.016). The analysis was then repeated using a dichotomized outcome, with respondents reporting heating “more than half the time” or “always” classified as practicing consistent thermal treatment and all others serving as the reference group. This threshold was selected because infrequent heating still results in regular consumption of unheated raw milk. Among all raw milk consumers (Model T_Any_), education level and nutrition beliefs were the only predictors that met the univariable screening threshold (p <0.10) and were carried forward to multivariable analysis. Subsequently, they were retained in the final multivariable model. Compared to respondents with a high school education or less, those with graduate or professional training had substantially higher odds of consistent thermal treatment (aOR: 13.87, 95% CI: 3.86–68.83), while differences for college graduates and those with some college or technical training were not significant. The proportion of respondents reporting consistent thermal treatment increased with education, from 32% among those with a high school education or less to 83% among those with graduate or professional training, and no respondent in the highest education group reported never heating raw milk. Respondents who were uncertain about the relative nutritional value of raw and pasteurized milk had lower odds of consistent thermal treatment compared to those who believed the two were nutritionally equivalent (aOR: 0.07, 95% CI: 0.01–0.36), although this estimate was based on a small group in which only 2 of 17 respondents reported consistent thermal treatment. Beliefs that either milk type was nutritionally superior were not significantly associated with heating behavior. This model demonstrated acceptable discrimination (AUC = 0.724) and adequate calibration (Hosmer-Lemeshow χ² = 1.90, df = 3, p = 0.594). It should be noted that the education association was driven largely by a small group of graduate-educated respondents (n = 23), and the nutrition beliefs association reflected lower odds of heating among respondents who expressed uncertainty (n = 17) rather than any belief. These findings should therefore be interpreted with caution. Among exclusive raw milk consumers (Model T_Exclusive_, n = 55), only political orientation met the screening threshold (p<0.1), and Republican orientation was not significantly associated with thermal treatment compared to Democrat orientation (aOR: 2.91, 95% CI: 0.88–10.28). This model showed weaker discrimination (AUC = 0.627); the Hosmer-Lemeshow test was not applicable, as a model containing a single categorical predictor yields fewer distinct fitted probabilities than the number of groups required for the test.

The dichotomized univariable analysis was repeated under two alternative definitions: (i) heating at least half the time versus less than half, and (ii) any heating versus never. The association with nutrition beliefs was stable in direction and magnitude across all three definitions (uncertain vs. nutritionally equivalent: aOR 0.07–0.11, all p ≤ 0.005). The association between education and thermal treatment was consistent in direction across definitions; under both alternative definitions, all respondents with graduate or professional training were classified as thermally treating their raw milk, so an odds ratio could not be estimated for that group. The association with political orientation among exclusive consumers was inconsistent in direction and magnitude across definitions (aOR: 2.91, 1.44, and 0.56) and did not reach significance under any.

## Discussion

This nationally representative survey was designed to determine the prevalence of raw milk consumption in the US, the proportion of raw milk consumers who heat their milk prior to consumption and identify predictors of these behaviors. The main findings were: (1) raw milk consumption prevalence is higher than previous national estimates, (2) multivariable analyses showed that raw milk consumption was associated with younger age, favorable taste and safety perceptions, and reliance on informal information sources, (3) household vulnerability was one of the strongest predictors of raw milk consumption, and (4) at-home thermal treatment behaviors were fairly common among raw milk consumers.

### Higher prevalence of raw milk consumption

Among 1,075 eligible US adult respondents, 15% reported consuming raw milk within the past six months, substantially higher than previous national estimates. Using nationally representative FDA surveys from 2016 and 2019, Lando et al. estimated that 4.4% of US adults had consumed raw milk at least once in the previous 12 months, while only 2% consumed raw milk at least monthly and 1% at least weekly ^6^. The authors found no significant differences in raw milk consumption between the 2016 and 2019 surveys. In contrast, our estimate of 15% was observed using a shorter, six-month recall period captured in 2024. A shorter recall period would be expected to capture fewer occasional consumers than a 12-month period, making the substantially higher prevalence observed in our study particularly notable.

However, differences in survey design and sampling prevent these estimates from being direct evidence of an increase in raw milk consumption over time. The FDA surveys asked respondents a binary “yes/no” question about whether they had consumed any raw milk in the previous 12 months, explicitly defining raw milk as “milk that was not pasteurized” and noting that it is “often sold off-the-farm or door-to-door” ^6^. In contrast, our survey asked respondents to identify which type of dairy milk they had consumed during the previous six months (raw, pasteurized, both, or neither), defined dairy milk as including milk from cows, goats, and sheep, and did not provide additional contextual information about raw milk. Question framing, survey administration, sampling, and the explicit description of raw milk may have influenced responses. Future research using standardized survey methods is needed to confirm the extent of an increase in raw milk consumption and identify contributing factors.

The higher prevalence observed in our study may also reflect real changes in consumer behavior such as increased interest in or access to raw milk since previous surveys. Prior research has described increasing consumer interest in local and unprocessed foods and greater availability of unpasteurized dairy products ^6^. Legal access may also contribute to consumption; Lando et al. found that adults living in states where retail sale of raw milk was legal were more likely to report raw milk consumption. In accordance with this, nine of the ten states that amended relevant raw milk laws between 2012 and 2018 expanded legal access, with 27 states permitting raw milk sales by 2019 ^2, 6^. Given the substantial increase of foodborne disease risk associated with unpasteurized dairy products, even modest increases in raw milk consumption could have important public health implications. Specifically, Costard et al. projected that a doubling of unpasteurized milk and cheese consumption could increase outbreak-related illnesses by approximately 96% ^2^. Determining whether our higher prevalence estimate reflects a true increase in consumption is essential for assessing the current and future burden of raw milk– associated foodborne illness.

### Respondent age, perceived safety and taste, and reliance on informal information sources strongly influenced consumption behavior

Our survey found that raw milk consumption is strongly associated with younger age, consistent with prior studies which have similarly identified higher consumption among younger and middle-aged adults ^6, 9, 26^. Our results showed perceptions of taste and safety were also among the strongest predictors of raw milk consumption – individuals who believed raw milk tasted equal or better than pasteurized milk, or who perceived similar or lower health risks, had substantially higher odds of raw milk consumption. Previous work has similarly identified sensory characteristics and perceived healthfulness as key motivators of raw milk consumption ^22, 26, 27^. Notably, individuals who perceived raw and pasteurized milk as equally risky also had significantly higher odds of raw milk consumption, suggesting that personal perception of the risk (or lack of it) may be a key driver of behavior. Similarly, Markham et al. found that raw milk consumers believed raw milk was safe and disagreed with government warnings ^22^. Finally, our finding that taste was one of the strongest predictors of raw milk consumption is consistent with studies reporting that consumers prefer the flavor of raw milk cheeses and perceive products labeled as “raw” to be of higher quality ^28^. Together, these findings suggest that consumption is strongly shaped by perceptions of raw milk safety and sensory benefits, calling for more targeted messaging to address these factors. Compared with general food safety instruction, pathogen-specific education has been shown to improve food safety knowledge and reduce high-risk food consumption, including raw milk, particularly among high-risk populations ^29^. Communication strategies that focus on general risk warnings may be insufficient to change behavior among consumers who already perceive raw milk as safe or desirable.

This point ties directly into respondents’ trusted information sources, which were strongly associated with raw milk consumption behavior. Individuals who relied on social media influencers, friends and family, or farmers were significantly more likely to consume raw milk compared to those who relied on medical professionals, highlighting the central role of informal information networks in health-related behaviors. Reliance on informal sources may also make it more difficult for traditional public health messaging, including pathogen-specific food safety education, to reach raw milk consumers. Thus, effective risk communication may require close consideration of the sources and channels through which consumers receive food safety information. Prior work has demonstrated that online and interpersonal sources often shape food safety beliefs more strongly than expert guidance, with consumers relying on these sources when forming perceptions of risk ^26^. Online food-related information is often evaluated through rapid, heuristic processes influenced by visual presentation and perceived relevance, rather than careful assessment of evidence or expertise ^30^. As a result, narratives that are simplified, emotionally engaging, or personally relevant may disproportionately influence beliefs about food and health. Specifically, social media can shape nutritional beliefs and diets, with such narratives amplified through these platforms and influencing food choices ^11^. Younger individuals may be disproportionately exposed to these forms of content, potentially contributing to observed age-related differences in consumption patterns ^11^.

In summary, raw milk consumers are not a homogeneous population but instead differ in their motivations, trusted information sources, and food safety knowledge. This aligns with prior work proposing distinct “mental models” of raw milk consumers, suggesting that understanding these differing perspectives may help inform more effective food safety education and risk communication strategies ^31^. Future research should prioritize understanding consumers’ motivations for consuming raw milk as a necessary first step toward delivering effective food safety education and risk communication.

### Living with high-risk individuals for foodborne infections was one of the strongest predictors of raw milk consumption

Among the 1,075 eligible respondents, 19% reported that at least one household member with high risk for foodborne infection had consumed raw milk in the past six months, including children ≤5 years old, adults ≥65 years old, immunocompromised individuals, or pregnant individuals. Children ≤5 years old (10% of all respondents) and adults ≥65 years old (9%) were the most frequently reported high-risk household members consuming raw milk. Among respondents who consumed raw milk themselves, approximately half reported living with a high-risk household member who also consumed raw milk, compared with 12% of respondents who consumed pasteurized milk only. Consistent with this pattern, living with a high-risk individual who consumed raw milk was one of the strongest and most consistent predictors of respondents’ own raw milk consumption across analyses. These findings suggest that raw milk consumption may cluster within households, including households in which individuals at increased risk of severe foodborne illness consume raw milk. This is particularly concerning given that children have been disproportionately represented among reported raw milk-associated illnesses, and both young children and older adults are at increased risk of severe illness following exposure to raw milk pathogens ^32^. Recent outbreak underscore these risks, including a 2026 multistate *E. coli* outbreak linked to raw milk cheese in which more than half of reported cases occurred among children aged five years or younger ^33^.

As a possible explanation of our findings, households with children or immunocompromised members may be more likely to seek foods they perceive as health-promoting, including raw milk, which is often viewed as more natural, less processed, and beneficial for immunity, allergies, or gut health ^34^. This explanation is consistent with studies suggesting that vulnerability may lead people to rely on heuristics rather than more central processing routes of information credibility ^35^. For example, parents of children with cancer were more likely to seek out health information but also more likely to believe COVID-19–related misinformation, a pattern attributed to increased stress and greater reliance on heuristic processing ^36^. Behavioral frameworks further suggest that heightened perceived concern for health may drive individuals to seek actions that reduce fear, including endorsement of prevention-focused beliefs ^35–37^.

However, our survey cannot determine why raw milk consumption clustered within these households; we did not include sufficient follow-up questions to characterize this association, limiting our ability to distinguish motivation from shared household behavior and exposure. Future work should inquire about presence of an at-risk household member in general, and subsequently their raw milk consumption, as well as assess raw milk consumption directly among high-risk individuals for foodborne infections.

### Thermal treatment behaviors were common, although predictors of this behavior were not consistent across analytic approaches

Thermal treatment of raw milk, which may represent an at-home intervention to reduce pathogen risk, was reported more than half the time or always by 47% of respondents consuming both raw and pasteurized milk and 38% of exclusive raw milk consumers. Although the effectiveness of reported thermal treatment practices could not be evaluated, approximately 4 in 10 respondents reported heating raw milk more than half the time or always. This novel finding suggests that US raw milk consumers may be receptive to interventions promoting thermal treatment as a food safety measure, a behavior studied in countries such as Italy and Ethiopia ^17–19^. Additionally, this finding has implications for future food safety risk assessments; existing assessments incorporate consumer behaviors such as storage time and temperature but generally assume that raw milk is consumed without prior household heat treatment ^38^. Consequently, failure to account for at-home thermal treatment could bias estimates of pathogen exposure among US raw milk consumers and overestimate the risk of foodborne disease. Future research should assess the effectiveness of at-home thermal treatment of raw milk, including the effects of heating temperature, duration, and storage time before heating.

Education showed a potential association with at-home thermal treatment among all raw milk consumers, although this finding was not consistent across analytic approaches. When heating frequency was examined using the original five-level response, education and perceived taste showed the strongest associations. When thermal treatment was dichotomized, respondents with graduate or professional training had significantly higher odds of consistent thermal treatment than those with a high school education or less. However, this association was driven largely by a small group of graduate-educated respondents (n = 23) and was not observed among exclusive raw milk consumers. Overall, these findings suggest a possible relationship between education and at-home thermal treatment, but larger studies are needed to determine reproducibility and identify other factors related to at-home thermal treatment among raw milk consumers.

### Limitations

While this survey provides novel insight into US raw milk consumption and motivation patterns, there exist certain limitations. Firstly, the prevalence report relies upon the assumption that our sample is representative of the target population. While we confirmed national representativeness by comparing a series of demographic variables with the US census, it is possible that there exist other confounders that would compromise the internal validity of our study. The QRS panel may introduce selection bias toward individuals willing to participate in online surveys, and in general, self-reported data introduces issues of selection and response biases. Further, individuals with stronger opinions about raw milk may have been more likely to complete the survey, introducing self-selection bias that could influence prevalence estimates and observed associations in either direction.

Additionally, while we compared raw milk to pasteurized in survey wording, it is possible that some respondents were unclear of the official definition leading to misclassification. If non-differential, this misclassification would likely bias results toward the null; however, if differential across respondent groups (e.g., by age or beliefs), the direction of bias is uncertain. It is also possible that respondents who in Q1 reported drinking only pasteurized milk in the past 6 months answered the question comparing raw and pasteurized milk taste (Q5) based on assumptions but may not have ever been in the presence of raw milk. This could introduce response bias and limit interpretation of perceived sensory factors impacting consumption behavior. Furthermore, this survey quantifies at-home heat treatment of raw milk, a previously under-characterized and potentially important risk mitigation measure, but respondents may have interpreted “heating” differently. This misconception could prompt wider confidence intervals due to a smaller sample size and therefore less power, biasing results towards the null, or potentially introducing some nondifferential misclassification. Finally, there could have been residual confounding from unmeasured variables (*e.g*., cultural or dietary patterns) that may have influenced variable associations through missing data.

## Conclusion

Our study reports that raw milk consumption frequency among US adults has considerably increased in recent years. Why cannot be definitively stated; our findings suggest that engaging in raw milk consumption is influenced by an interplay of sensory perception, household context, and risk interpretation, rather than a single dominant factor. Further, the quantification of consumers heating their milk at home represents a previously under-characterized and important risk mitigation measure. Thermal treatment behaviors were not found to be an extension of consumption patterns but were rather shaped by distinct belief systems and sociopolitical factors. Our insights will be valuable in informing policymakers about the extent and potential modifiers of US raw milk consumption.

## Acknowledgements

This research was supported by the Einhorn Discovery Grant and Cornell College of Arts and Sciences Undergraduate Research Fund to SR; the U.S. Department of Agriculture (USDA) National Institute of Food and Agriculture (NIFA) Hatch project 7010559 to RI; and the National Institutes of Health (NIH) Office of the Director under training grant T32OD011000 to KJK. The content is solely the responsibility of the authors and does not necessarily represent the official views of the funding agencies.

## Declarations

### Ethics approval and consent to participate

The Cornell University Institutional Review Board determined this study to be exempt (IRB #IRB0149147).

## Data availability

All data and analysis code generated or used during this study are publicly available in the GitHub repository at https://github.com/IvanekLab/Raw-Milk-Survey-Data-Analysis.

## Tables + Figures

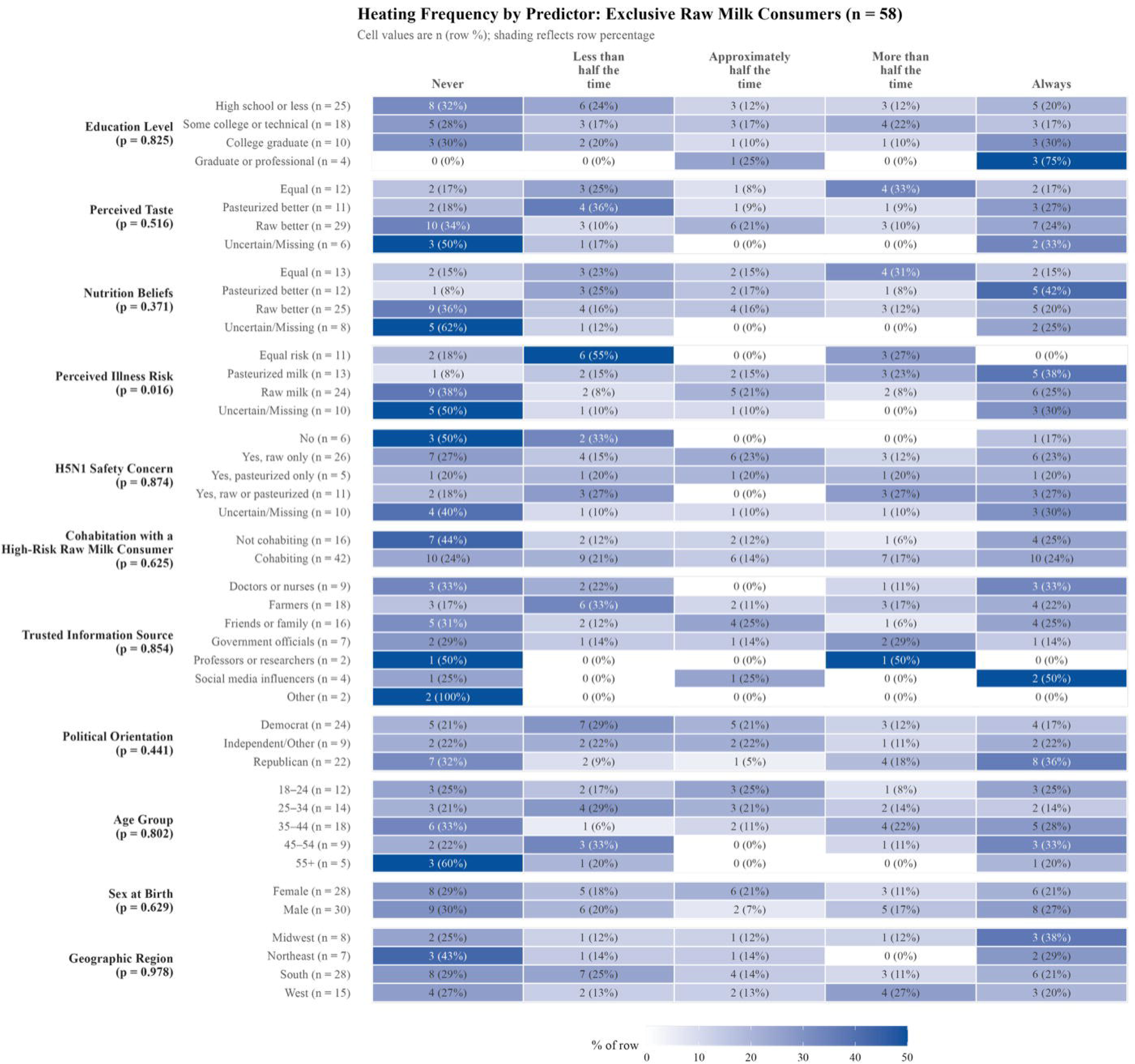

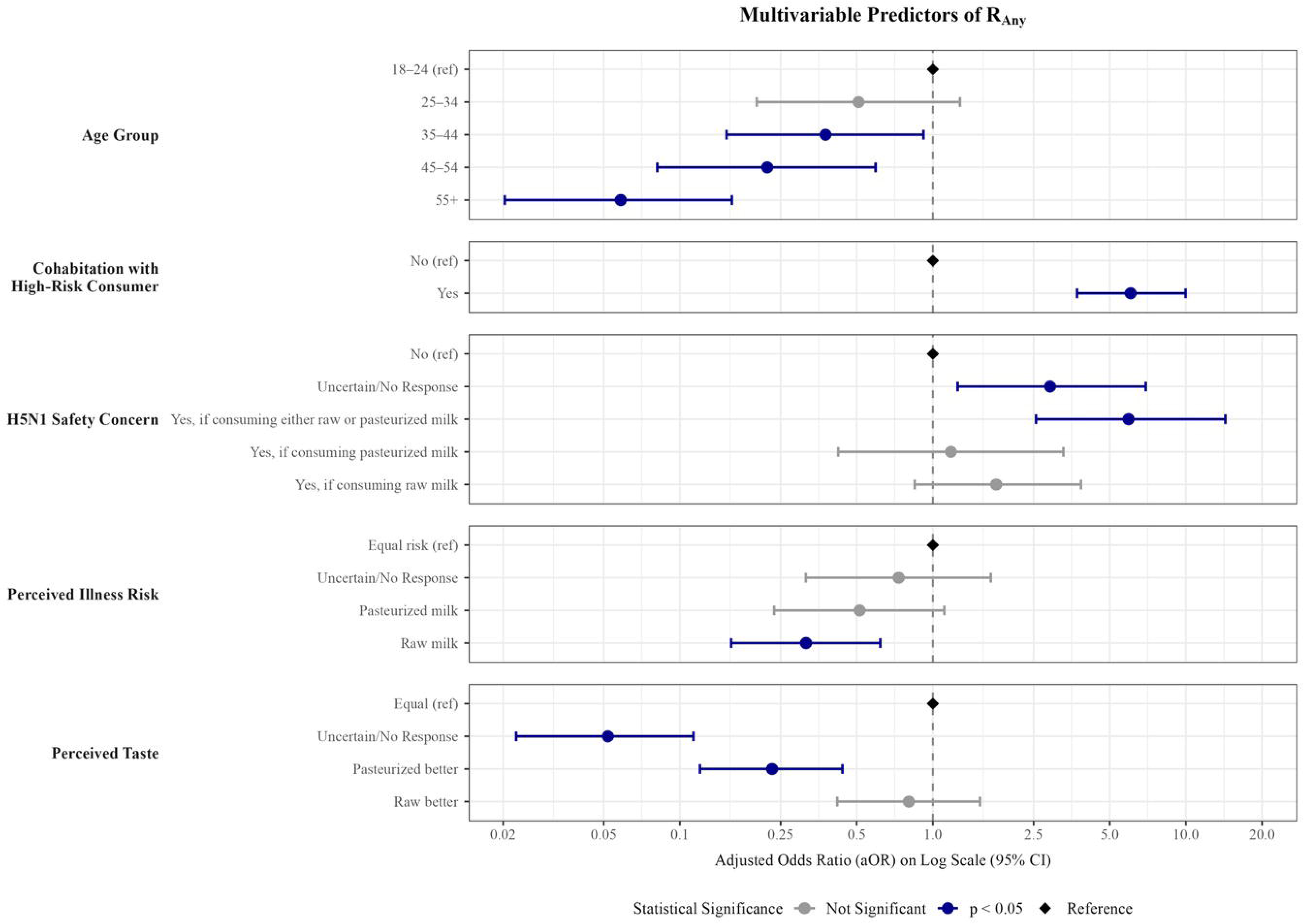

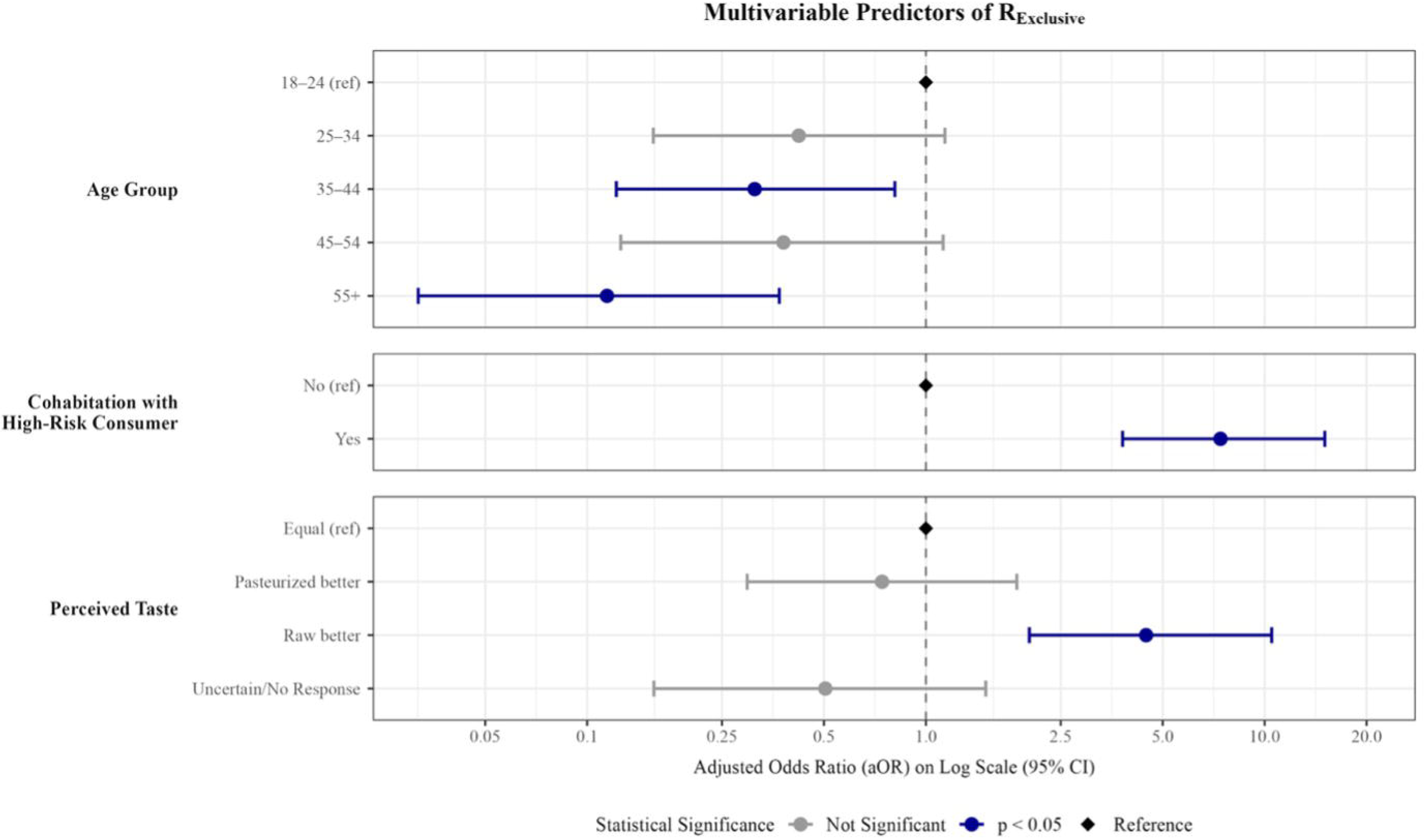

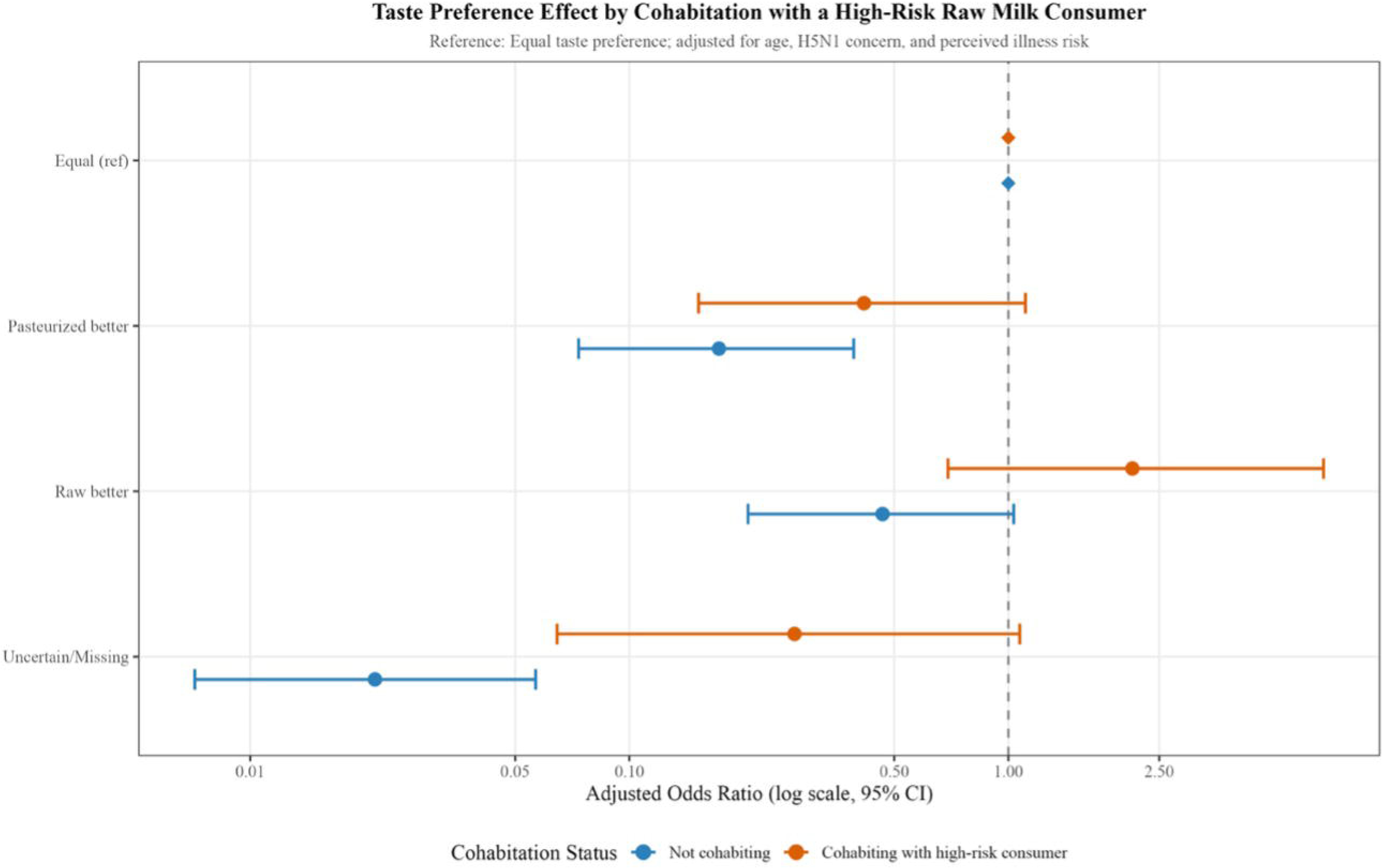

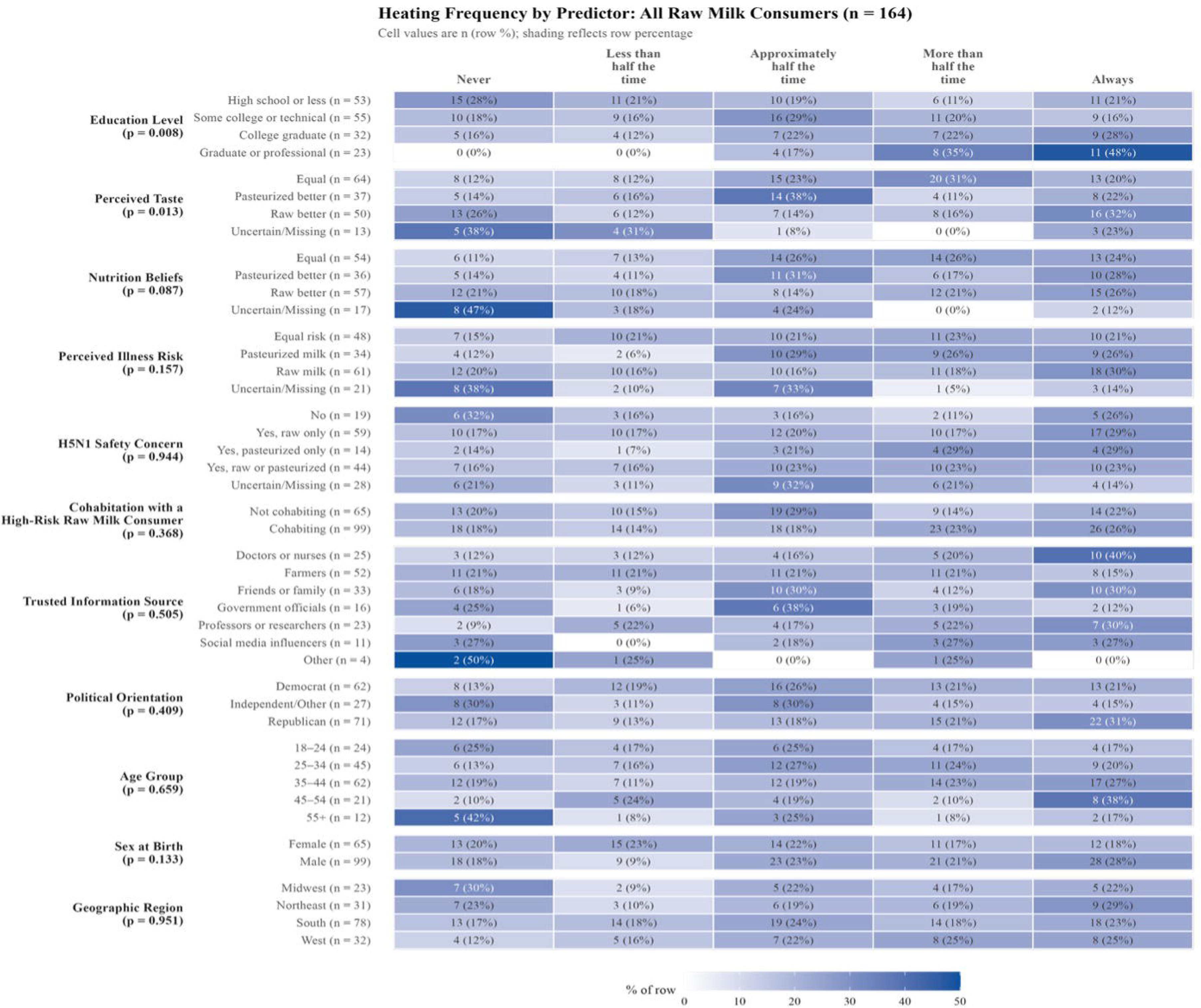

## References

1. U.S. Food and Drug Administration. Raw Milk Misconceptions and the Danger of Raw Milk Consumption: U.S. Food and Drug Administration; [Available from: https://www.fda.gov/food/buy-store-serve-safe-food/raw-milk-misconceptions-and-danger-raw-milk-consumption?

2. Costard S, Espejo L, Groenendaal H, Zagmutt FJ. Outbreak-Related Disease Burden Associated with Consumption of Unpasteurized Cow’s Milk and Cheese, United States, 2009-2014. Emerg Infect Dis. 2017;23(6):957–64.

3. Ablan M, Canning M, Koski L, Landsman L, Stapleton GS, Nichols M, et al. Responding to outbreaks of illness linked to unpasteurized milk: A needs assessment of state health and agriculture departments. Zoonoses Public Health. 2024;71(5):480–8.

4. Koski L, Kisselburgh H, Landsman L, Hulkower R, Howard-Williams M, Salah Z, et al. Foodborne illness outbreaks linked to unpasteurised milk and relationship to changes in state laws – United States, 1998-2018. Epidemiol Infect. 2022;150:e183.

5. David SD. Raw milk in court: implications for public health policy and practice. Public Health Rep. 2012;127(6):598–601.

6. Lando AM, Bazaco MC, Parker CC, Ferguson M. Characteristics of U.S. Consumers Reporting Past Year Intake of Raw (Unpasteurized) Milk: Results from the 2016 Food Safety Survey and 2019 Food Safety and Nutrition Survey. J Food Prot. 2022;85(7):1036–43.

7. Rhodes MT, Kuchler F, McClelland K, Hamrick KS. Consumer Food Safety Practices: Raw Milk Consumption and Food Thermometer Use. Washington, DC: U.S. Department of Agriculture, Economic Research Service; 2019 January 2019.

8. Lucey JA. Raw Milk Consumption: Risks and Benefits. Nutr Today. 2015;50(4):189–93.

9. Bigouette JP, Bethel JW, Bovbjerg ML, Waite-Cusic JG, Häse CC, Poulsen KP. Knowledge, Attitudes and Practices Regarding Raw Milk Consumption in the Pacific Northwest. Food Protection Trends. 2018;38(2):104–10.

10. Knutson RD, Currier RW, Ribera L, Goeringer P. Asymmetry in Raw Milk Safety Perceptions and Information: Implications for Risk in Fresh Produce Marketing and Policy. 1st Joint EAAE/AAEA Seminar “The Economics of Food, Food Choice and Health”; September 15–17, 2010; Freising, Germany2010.

11. Capocasa M, Venier D, Venier B, Chiarla C. Exploring nutritional myths and fake news: Impact and counteractions. Clin Nutr ESPEN. 2026;72:102947.

12. Román S, Sánchez-Siles LM, Siegrist M. The importance of food naturalness for consumers: Results of a systematic review. Trends in Food Science & Technology. 2017;67:44–57.

13. U.S. Food and Drug Administration. Pasteurized Milk Ordinance Centennial: U.S. Food and Drug Administration; [Available from: https://www.fda.gov/food/milk-guidance-documents-regulatory-information/pasteurized-milk-ordinance-centennial.

14. Wisconsin Department of Agriculture TaCP. Raw Milk: Wisconsin Department of Agriculture, Trade and Consumer Protection; [Available from: https://datcp.wi.gov/Pages/Programs_Services/RawMilk.aspx.

15. Wittwer M, Hammer P, Runge M, Valentin-Weigand P, Neubauer H, Henning K, et al. Inactivation Kinetics of Coxiella burnetii During High-Temperature Short-Time Pasteurization of Milk. Frontiers in Microbiology. 2022;Volume 12 – 2021.

16. International Dairy Foods Association. Pasteurization: International Dairy Foods Association; [Available from: https://www.idfa.org/pasteurization.

17. Pannella G, Messia MC, Tremonte P, Tipaldi L, La Gatta B, Lombardi SJ, et al. Concerns and solutions for raw milk from vending machines. Journal of Food Processing and Preservation. 2019;43(10):e14140.

18. Duguma B. Milk composition, traditional processing, marketing, and consumption among smallholder dairy farmers in selected towns of Jimma Zone, Oromia Regional State, Ethiopia. Food Sci Nutr. 2022;10(9):2879–95.

19. Amenu K, Wieland B, Szonyi B, Grace D. Milk handling practices and consumption behavior among Borana pastoralists in southern Ethiopia. J Health Popul Nutr. 2019;38(1):6.

20. Hartschuh J. Food Preservation: Pasteurization of Raw Milk for Home Consumption: Ohio State University Extension; 2025 [Available from: https://cfaes.osu.edu/fact-sheet/food-preservation-pasteurization-raw-milk-home-consumption.

21. Arnold L, Gustafson CJ. Home Pasteurization of Milk. American Journal of Public Health and the Nations Health. 1930;20(10):1065–70.

22. Markham L, Auld G, Bunning M, Thilmany D. Attitudes and Beliefs of Raw Milk Consumers in Northern Colorado. Journal of Hunger & Environmental Nutrition. 2014;9(4):546– 64.

23. Qualtrics. Online Research Panels & Samples for Surveys 2025 [Available from: https://www.qualtrics.com/research-services/online-sample/.

24. U.S. Census Bureau. Census Regions and Divisions of the United States: U.S. Census Bureau; [Available from: https://www2.census.gov/geo/pdfs/maps-data/maps/reference/us_regdiv.pdf.

25. U.S. Census Bureau. 2020 Census Redistricting Data [PL 94-171]: U.S. Department of Commerce; 2021 [Available from: data.census.gov.

26. Headrick ML, Timbo B, Klontz KC, Werner SB. Profile of raw milk consumers in California. Public Health Rep. 1997;112(5):418–22.

27. Mullin GE, Belkoff SM. Survey to determine why people drink raw milk. Glob Adv Health Med. 2014;3(6):19–24.

28. Colonna A, Durham C, Meunier-Goddik L. Factors affecting consumers’ preferences for and purchasing decisions regarding pasteurized and raw milk specialty cheeses. J Dairy Sci. 2011;94(10):5217–26.

29. Kendall P, Scharff R, Baker S, LeJeune J, Sofos J, Medeiros L. Food Safety Instruction Improves Knowledge and Behavior Risk and Protection Factors for Foodborne Illnesses in Pregnant Populations. Matern Child Health J. 2017;21(8):1686–98.

30. Sillence E, Hardy C, Medeiros LC, LeJeune JT. Examining trust factors in online food risk information: The case of unpasteurized or ‘raw’ milk. Appetite. 2016;99:200–10.

31. Medeiros LC, LeJeune J. Why Do Consumers Drink Unpasteurized Milk? A Preliminary Mental Model. Food Protection Trends. 2016;36(6):428–42.

32. Robinson TJ, Scheftel JM, Smith KE. Raw milk consumption among patients with non-outbreak-related enteric infections, Minnesota, USA, 2001-2010. Emerg Infect Dis. 2014;20(1):38–44.

33. Control CfD, Prevention. Investigation Update: E. coli Outbreak, September 2026: Centers for Disease Control and Prevention; 2026 [updated September 25, 2026. Available from: https://www.cdc.gov/ecoli/outbreaks/raw-milk-cheese-09-26/investigation.html.

34. Zaitlin P, Dwyer J, Gleason G. Mistaken Beliefs and the Facts About Milk and Dairy Foods. Nutrition Today. 2013;48(3):135–43.

35. Cameron KA. A practitioner’s guide to persuasion: an overview of 15 selected persuasion theories, models and frameworks. Patient Educ Couns. 2009;74(3):309–17.

36. Guidry JPD, Miller CA, Ksinan AJ, Rohan JM, Winter MA, Carlyle KE, et al. COVID-19-Related Misinformation among Parents of Patients with Pediatric Cancer. Emerg Infect Dis. 2021;27(2):650–2.

37. Witte K. Putting the fear back into fear appeals: The extended parallel process model. Communication Monographs. 1992;59(4):329–49.

38. Crotta M, Paterlini F, Rizzi R, Guitian J. Consumers’ behavior in quantitative microbial risk assessment for pathogens in raw milk: Incorporation of the likelihood of consumption as a function of storage time and temperature. J Dairy Sci. 2016;99(2):1029–38.

