## Supplementary Materials for "Why Americans Choose Raw Milk: Evidence from a National Quantitative Survey"

**Supplementary Table S1**. Survey questions, answer choices, and variables
Answers to all questions are mandatory

| Question (Q) | Answer Choices | Variable |
|---|---|---|
| (Q9) What is your age? | Under 18; 18-24; 25-34; 35-44; 45-54; 55-64; 65 or over; Prefer not to answer | Potential confounder |
| (Q11) In which state do you currently reside? | [Drop down menu; 50 continental states unless Qualtrics includes Puerto Rico + Minor Outlying Islands]; Prefer not to answer | Potential confounder |
| (Q1) What dairy milk have you consumed in the last 6 months? This means milk from animals such as cows, goats, or sheep. | Raw dairy milk; Pasteurized dairy milk; Both raw and pasteurized dairy milk; Neither raw nor pasteurized dairy milk; Prefer not to answer | Primary outcome |
| Presented only to respondents who in Q1 responded “Raw dairy milk” or “Both raw and pasteurized dairy milk”: (Q2) How frequently do you heat up raw milk (such as microwave, boiling, or steaming) before consumption? | Never; Less than half the time; Approximately half the time; More than half the time; Always; Prefer not to answer | Secondary outcome |
| (Q3) If you had a question about the safety of milk, which source of information would you trust the most? | Friends or family; Farmers; Government officials; Social media influencers; Professors or researchers; Doctors or nurses; Healer, chiropractor, naturopath; Other [specify]; Prefer not to answer | Predictor |
| (Q4) Who in your household has consumed raw dairy milk in the last 6 months? Select all that apply. | Children aged 5 or younger; Adults aged 65 or older; People with limited immunity (e.g., getting chemotherapy); Pregnant individuals; None of the above groups apply; Prefer not to answer | Predictor |
| (Q5) Based on what you know, how does the taste of raw milk compare to pasteurized milk? | Raw is better; Pasteurized is better; They are equal; I don’t know; Prefer not to answer | Predictor |
| (Q6) Based on what you know, how does the nutritional value of raw milk compare to pasteurized milk? | Raw is better; Pasteurized is better; They are equal; I don’t know; Prefer not to answer | Predictor |
| (Q7) Based on what you know, which milk is more likely to make someone sick? | Raw milk; Pasteurized milk; They are equal; I don’t know; Prefer not to answer | Predictor |

| | | |
|---|---|---|
| (Q8) Based on what you know, should consumers worry about milk safety during a bird flu (H5N1) outbreak in dairy cattle? | No; Yes, if consuming raw milk; Yes, if consuming pasteurized milk; Yes, if consuming either raw or pasteurized milk; I don't know; Prefer not to answer | Predictor |
| (Q10) What is your sex assigned at birth? | Male; Female; Other [specify?]; Prefer not to answer | Potential confounder |
| (Q12) What is your highest level of education? | Below high school; Some high school; High school graduate (grade 12 or GED certificate); Technical, trade, or vocational school after high school; Some college or 2 year Associate Degree; College graduate (BS, BA, or other 4-year degree); Post-graduate training or professional schooling after college (e.g. toward a Master's Degree, PhD, or law/medical degree); Prefer not to answer | Potential confounder |
| (Q13) Generally speaking, when it comes to political parties in the United States, how would you best describe yourself? | Strong Democrat; Not very strong Democrat; Independent, close to Democrat; Independent (close to neither); Independent, close to Republican; Not very strong Republican; Strong Republican; Other party affiliation [specify]; Prefer not to answer | Potential confounder |

**Supplementary Table S2.** Analytic samples for each model

| Model | Sample sub-population | Full analytical sample | Outcome comparison | n in the final model |
|---|---|---|---|---|
| $R_{Any}$ | Respondents who reported consuming dairy milk | n = 898 | 164 both exclusive raw milk consumers and consumers of both milk types vs. 734 pasteurized-only consumers | n = 898 |
| $R_{Exclusive}$ | Respondents who reported consuming dairy milk | n = 898 | 58 exclusive raw milk consumers vs. 840 consumers of both milk types and of pasteurized milk only | n = 898 |
| $T_{Any}$ | Consumers of both milk types and raw milk only | n = 164 | 72 consistent heaters vs. 92 heating half the time or less | n = 163 |
| $T_{Exclusive}$ | Exclusive raw milk consumers | n = 58 | 22 consistent heaters vs. 36 heating less than half the time | n = 55 |

Note: The 164 raw milk consumers comprise 58 who consumed raw milk exclusively and 106 who consumed both raw and pasteurized milk. Model n differs from the sample size where respondents had missing values on a retained predictor: one respondent was excluded from Model $T_{Any}$ for missing education, and three from Model $T_{Exclusive}$ for missing political orientation.

**Supplementary Text S1**. Qualtrics Research Services

The sample of participants was recruited through Qualtrics Research Services (QRS) sampling, which displays this research opportunity from a flyer, advertisement, email/phone call, presentation, or similar prospective recruitment strategy [23]. The majority of QRS samples come from traditional, actively managed, double-opt-in market research panels [23]. Panel members are sent an email invitation or prompted on the respective survey platform to proceed with a given survey. The typical survey invitation sent by QRS is generally very simple and generic. It provides a hyperlink which will take the respondent to the survey. Often, potential respondents are sent an email invitation informing them that the survey is for research purposes only, how long the survey is expected to take, and what incentives are available. To avoid self-selection bias, survey invitations do not include specific details about the contents of the survey and are instead kept very general.

**Supplementary Text S2.** Interaction Term in Model $R_{Any}$

A two-way interaction between perceived taste and cohabitation with a high-risk raw milk consumer was evaluated in Model $R_{Any}$. In the full interaction model, two of the three interaction terms reached individual significance (Uncertain/No response taste × cohabiting: ratio of odds ratios: p = 0.009; raw better × cohabiting: p = 0.019), while pasteurized better × cohabiting did not (p = 0.254). After stratifying the estimates by the cohabitation variable to better understand their individual effects, we observed that among respondents not cohabiting with a high-risk raw milk consumer, taste preference was strongly associated with consumption across all categories, whereas among those who were cohabiting, all taste effects were closer to the null and not statistically significant (Supplementary Figure S1).

However, this interaction was one of ten evaluated, and it did not remain significant after correction for multiple comparisons under either the Bonferroni or Benjamini-Hochberg method (raw p = 0.026; both adjusted p = 0.256). Additionally, the "Uncertain/No response" taste category differed substantially in composition between strata, comprising 55% of respondents not cohabiting with a high-risk consumer compared to 15% of those who were, which further complicates interpretation. Given these considerations and the small sample sizes in the compared groups, the interaction was not retained in the final model and is reported as an exploratory finding.

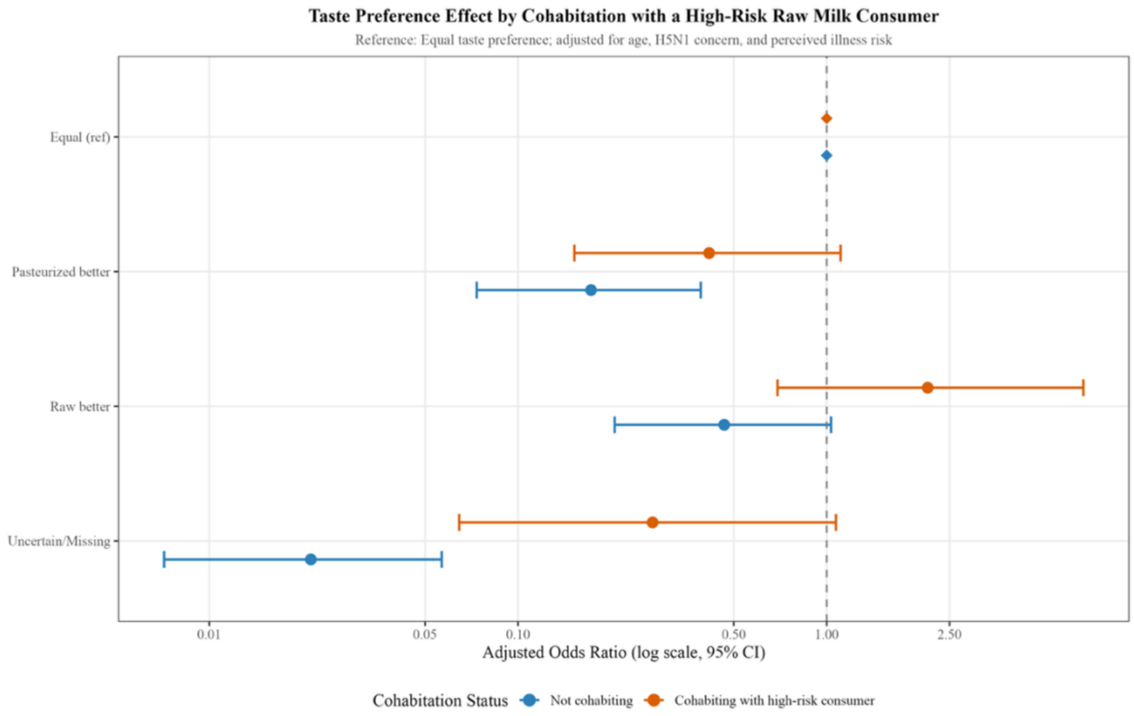


**Supplementary Figure S1.** Interaction between perceived taste and cohabitation with a high-risk individual who had consumed raw milk. Adjusted odds ratios (aORs) and 95% confidence intervals for any raw milk consumption according to perceived taste preference, stratified by cohabitation status. Respondents who perceived raw and pasteurized milk as tasting equal served as the reference category. Estimates were derived from Model $R_{Any}$, adjusted for age, H5N1 safety concern, and perceived illness risk; the dashed vertical line indicates the null value (aOR = 1).

**Supplementary Text S3.** Supplementary Figure S2 shows the heating frequency by predictor among exclusive consumers and consumers of raw and pasteurized milk (n = 164). Values are n (row %) across the five original response categories for at-home thermal treatment (Q2), prior to dichotomization. P-values are from Fisher's exact tests and assess the association between each predictor and the full five-level heating frequency response. Education level ($p = 0.008$) and perceived taste ($p = 0.013$) showed the strongest associations. Reference categories are listed first within each variable. Percentages are row percentages and may not sum to 100% due to rounding. Education displays a clear pattern; those with higher education were less likely to never heat their milk, and more likely to always heat it.

**Supplementary Figure S2.** Heatmap showing heating frequency by predictor in all raw milk consumers (n = 164)

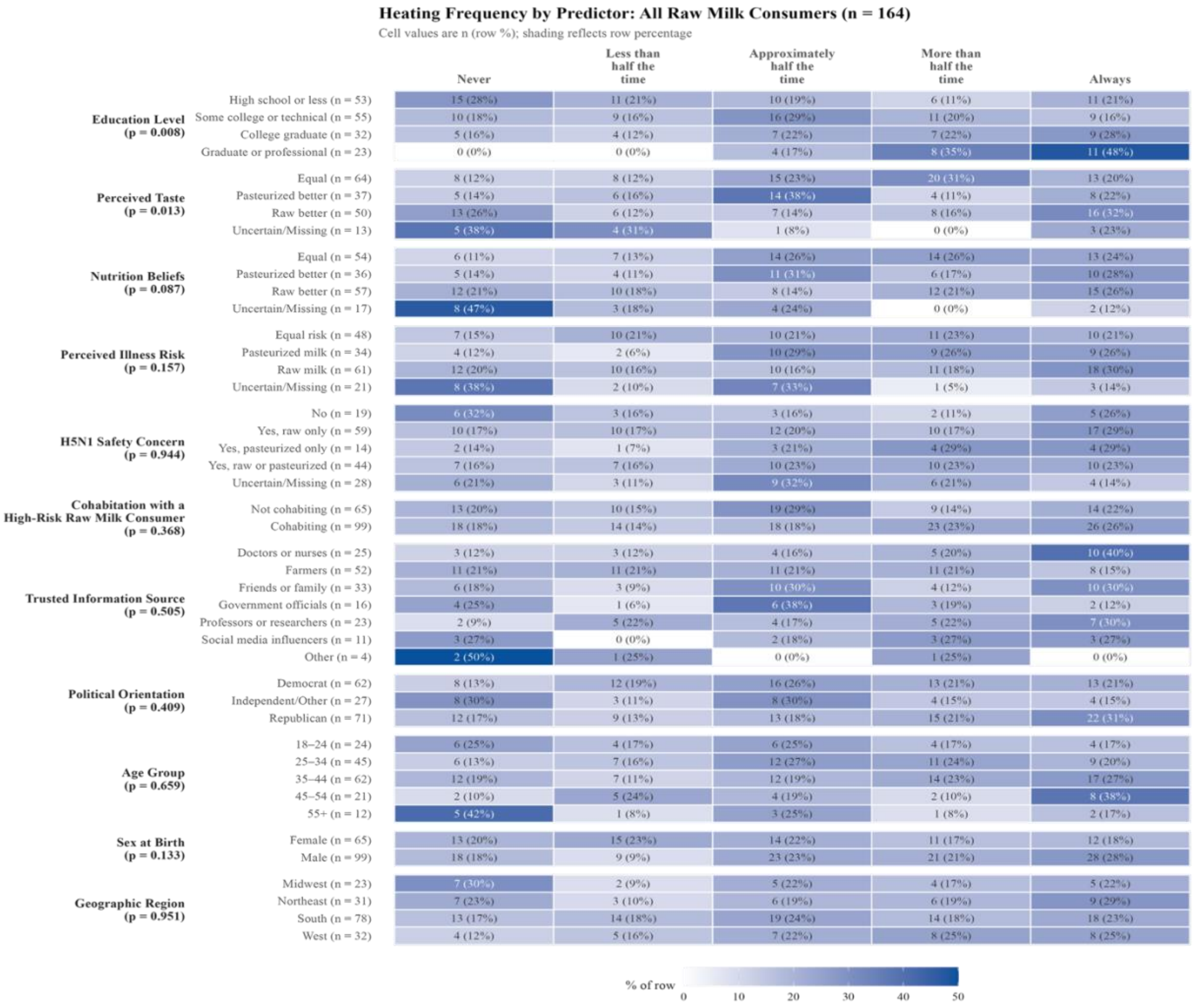


**Supplementary Text S4.** Supplementary Figure S3 shows the heating frequency by predictor among exclusive raw milk consumers (n = 58). Values are n (row %) across the five original response categories for at-home thermal treatment (Q2), prior to dichotomization. P-values are from Fisher's exact tests and assess the association between each predictor and the full heating frequency response. Perceived illness risk showed the strongest association ($p = 0.016$). Reference categories are listed first within each variable. Percentages are row percentages and may not sum to 100% due to rounding. Cell counts are small throughout, and estimates should be interpreted with caution.

**Supplementary Figure S3.** Heatmap showing heating frequency by predictor in exclusive raw milk consumers (n = 58)

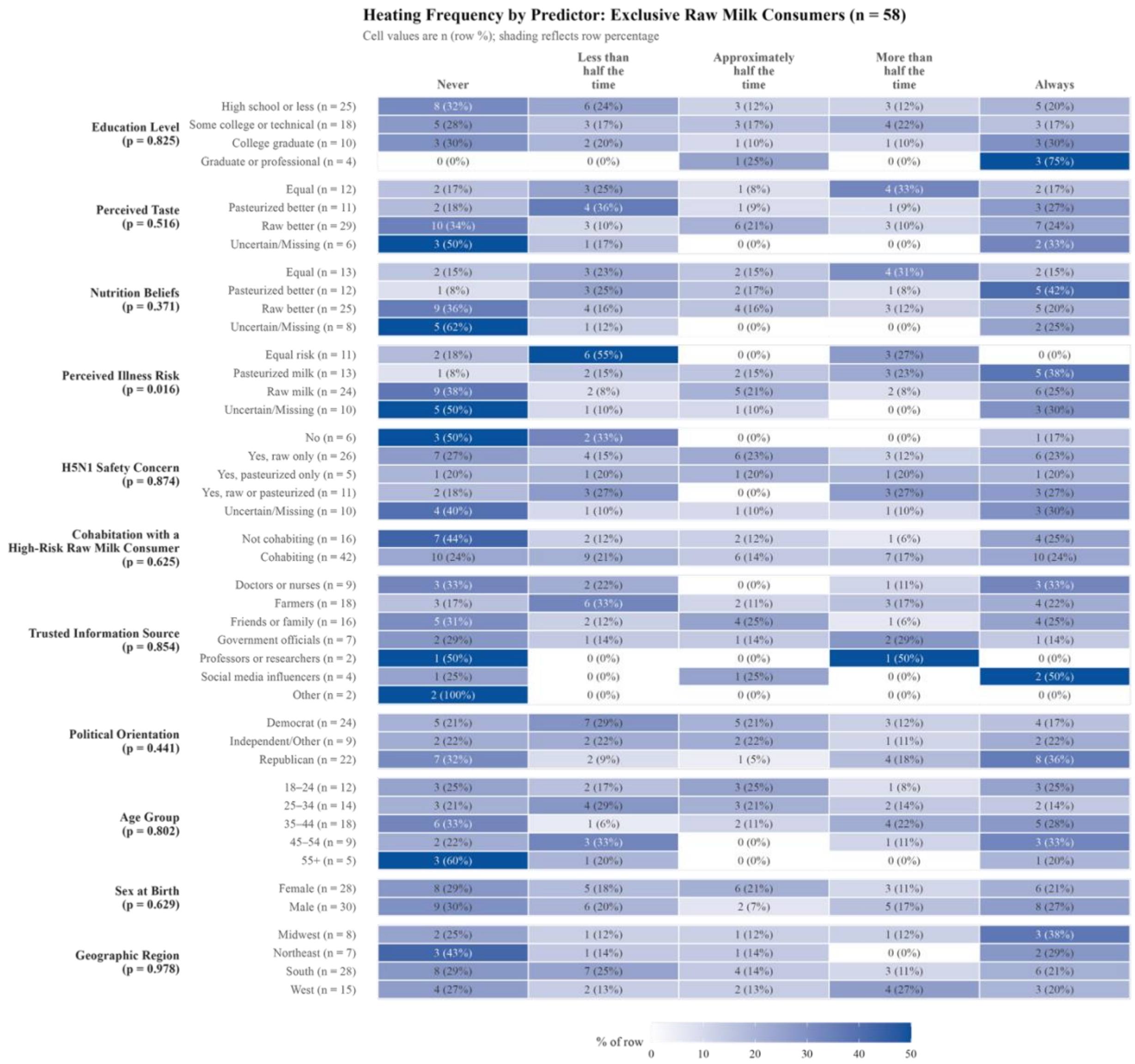
